# Sex and autism share cell type-specific effects on cortical gene expression

**DOI:** 10.64898/2026.10.28.26366315

**Authors:** Gabrielle Dugan, Alex R. DeCasien, Ramin Ali Marandi Ghoddousi, Rachel Smith, Siyuan Liu, Pavan Auluck, Stefano Marenco, Daniel Geschwind, Jason P. Lerch, Armin Raznahan

## Abstract

Potent sex effects on the prevalence and presentation of autism could theoretically reflect overlapping neurobiology, but tests in brain gene expression have so far been limited to bulk tissue — overlooking cell type heterogeneity. Here, we use newly available single-nucleus RNA-sequencing datasets, to test if regional, cell type-specific sex effects on gene expression overlap with those of autism (using schizophrenia as a comparison condition). Cellular gene expression effects were estimated for sex across six cortical regions (169 samples, 30 individuals), and for autism (63 individuals, 33 cases), and schizophrenia (140 individuals, 65 cases) in frontal cortex. Genes with significant sex effects showed a stronger overlap with those differentially expressed in autism than schizophrenia. Multivariate analyses revealed a significant shared axis of transcriptomic effects between sex and autism (but not schizophrenia). This axis featured a protein-protein interaction network enriched for autism risk genes and synaptic biology, downregulated in autism across cell types, and female biased in fusiform neurons, but male biased in angular gyrus glial cells. Thus, the autism transcriptomic signatures in frontal cortex show cell type- and region-specific alignments with sex effects on gene expression — pointing to a complexity in overlaps that informs theoretical models of sex modulation of autism liability.

## Introduction

Almost all neuropsychiatric conditions show sex differences in prevalence, presentation, and treatment response ^1–3^. For example, males show higher prevalence of early childhood conditions, such as autism spectrum conditions (henceforth autism) ^3–5^ and attention-deficit/hyperactivity disorder (ADHD) ^3,6,7^, whereas females show higher rates of adolescent-emergent anxiety and major depressive disorder ^8^. These observations may in part reflect intersections between the neurobiology of sex and neuropsychiatric conditions ^9^ — motivating study designs capable of testing for, quantifying and localizing such intersections, which remained to be assessed with cell type resolution in human tissue.

Autism is one of the most strongly sex-biased conditions, with a 3-to-5-fold greater prevalence in males than females ^10,11^ that is seen across diverse sociocultural contexts ^11–13^. There are also marked sex differences in the average profile of clinical features: females with autism show fewer restricted behaviors, more internalizing symptoms ^14–18^, and a higher likelihood for comorbid intellectual disability ^19,20^ than males. The differential fit of these presentations with autism diagnostic criteria originally established in male-biased cohorts ^21^ may lead to diagnostic biasing ^22–24^, yet different presentation in males and females itself represents a fundamental sex difference. Moreover, the male bias in autism prevalence is shared with several other phenotypically distinct neurodevelopmental disorders of early childhood onset ^25^ (e.g. ADHD ^7^, obsessive compulsive disorder ^26^, and disorders of language and motor coordination ^27^) — suggesting that there may be more fundamental intersections between the neurobiology of sex and these early emerging conditions, which is seen most strongly for autism.

Several studies have tested for overlaps between neurobiological correlates of sex and autism ^14,28,29^, although this literature has focused on neuroimaging data ^28,30–37^, with fewer reports considering measures of gene expression ^23,38–40^ that can be more directly integrated with known genetic risk factors for autism ^41^. The “female protective effects” model posits a higher liability threshold for autism in females^42^, with empirical support from greater etiological load in females with autism than males^43^. There is emerging evidence, from studies of bulk brain tissue, in support of this that there may be an overlap between female-biased genes enriched for synaptic annotations and genes that are downregulated in autism ^44^. However, the exclusive use of bulk tissue has meant this work is unable to account for a key new insight afforded by single-cell sequencing technology ^45,46^: that effects of sex and autism on gene expression are both cell type-specific. Further, previous work on transcriptomic overlaps between sex and autism has been limited to few brain regions, and it has recently been shown that sex differences on cell type gene expression can vary by brain region ^45^. Thus, there is a pressing need to reappraise transcriptomic overlaps between sex and autism at single-cell scale — in a manner that takes advantage of the latest multi-region single-cell surveys of sex effects on brain gene expression ^45^.

Here, we leverage newly available single-cell datasets from postmortem human cortical tissue to test for the overlapping effects of sex ^45^ and autism ^46^ on cell type-specific gene expression. We also perform a parallel single-cell analysis of transcriptomic overlaps between sex and schizophrenia ^47^ — a less sex-biased condition at early onset ^48,49^. We test for transcriptomic overlaps using multiple complementary analytic methods, which converge on the same core findings. First, sex differences in cortical gene expression overlap more strongly with gene expression changes in autism than in schizophrenia. Second, transcriptomic overlaps between sex and autism are most consistently prominent for excitatory and inhibitory neurons — especially for neuronal sex effects in the fusiform gyrus. Third, within these neuronal populations, the transcriptomic convergence between sex and autism highlights genes enriched for synaptic biology, which harbor genetic risk for autism, are downregulated in autism, and upregulated in females. These findings highlight a region- and cell type-varying version of the “female protective effect” model ^42,50^ for sex differences in autism — whereby females may possess a higher liability threshold for manifestation of autism through a female-bias of autism-relevant synaptic pathways in select neural cell types that is more prominent in some cortical regions than others.

## Results

### Datasets overview and analytical framework

We leveraged recently published single-cell datasets to test for and localize shared transcriptomic signatures between sex and autism across human brain cell types. As a comparison condition to benchmark the results, we performed a parallel analysis on sex and schizophrenia — which has a smaller sex bias than autism in early development (44).

Cell type-specific gene expression effects were estimated using available single-nucleus RNA-sequencing (snRNAseq) post-mortem data as follows: (i) for sex in 169 samples across 6 cortical regions [fusiform gyrus (FusGyr), inferior lateral temporal occipital cortex (InfLTO), caudal insula (CauIns), intraparietal sulcus (IntParSul), retrosplenial cortex (RetCor), angular gyrus (AngGyr)] from 30 individuals (15 male / 15 female) ^45^; (ii) for autism in frontal cortex from 63 individuals (33 cases / 30 controls) ^46^; and (iii) for schizophrenia in frontal cortex from 140 individuals (65 cases / 75 controls) ^47^ (**Fig 1A; Table S1**). From these datasets, we extracted differential gene expression statistics, using log fold-changes (logFC) and false discovery rate (FDR) adjusted p-values (*q*).

**Fig. 1:**
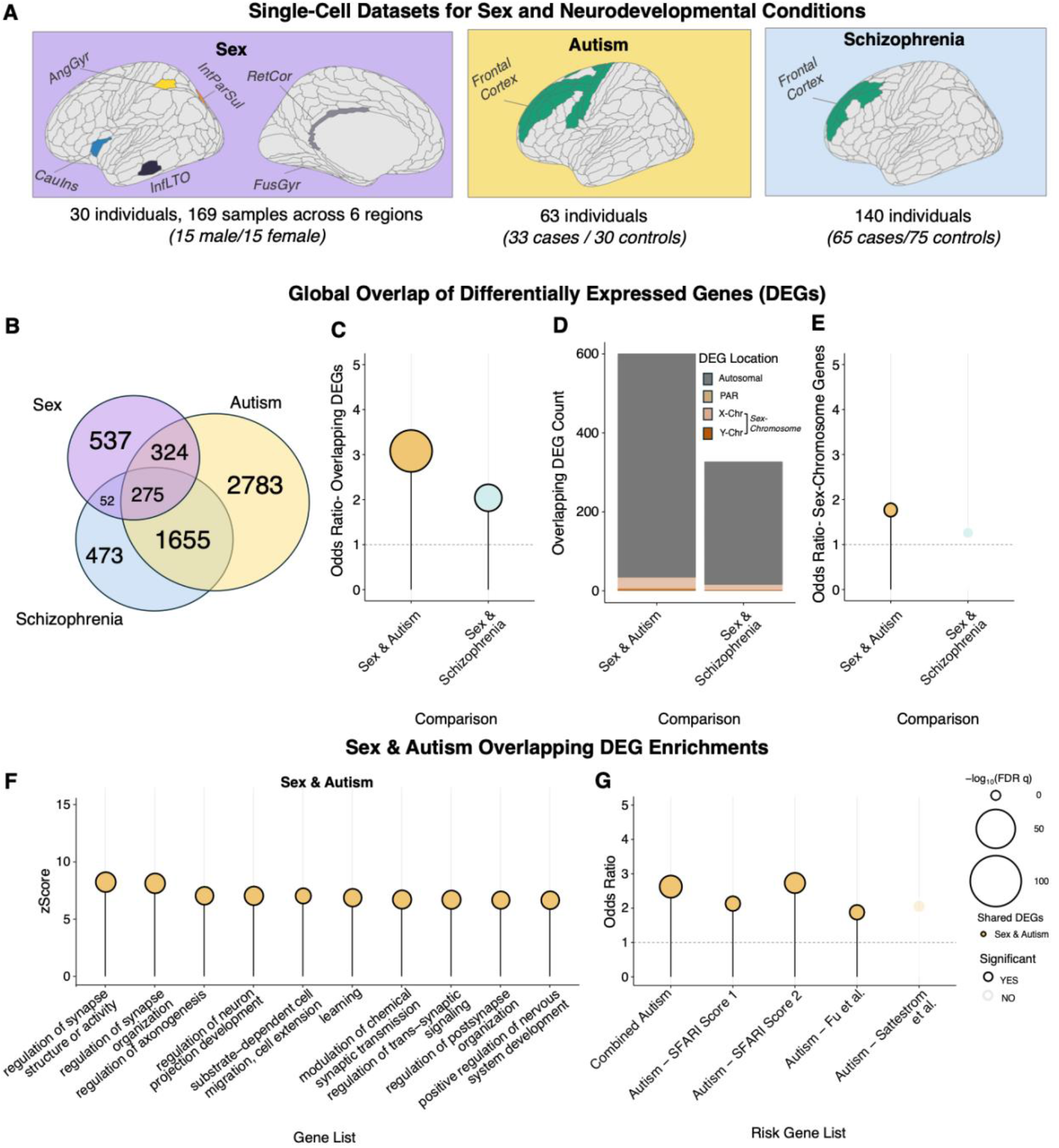
Overview of datasets and overlapping differentially expressed genes (DEGs) between datasets. **(A)** Schematic overview of sex (purple), autism (yellow), and schizophrenia (blue) datasets, highlighting regions used for single-nucleus RNA-sequencing (snRNAseq) analyses of differential expression **(B)** Venn diagram of overlaps between the global set of DEG (collapsing across cell types) in each of the 3 datasets **(C)** Statistical significance (Fisher’s exact test) of DEG overlaps between sex and autism (gold) and between sex and schizophrenia (blue) **(D)** Bar chart showing breakdown of overlapping DEGs according to chromosomal location: autosomal (dark grey), X-chromosome (pale orange), Y-chromosome (dark tan), and PAR genes (beige). **(E)** Lollipop plot showing odds ratio (OR) of sex-chromosome DEGs compared to background for sex and autism (gold) and sex and schizophrenia (blue). (**F)** Lollipop plot for Gene Ontology (GO) ^62^ enrichment of shared DEGs between sex and autism from the biological process (BP) ontology, generated using clusterProfiler ^63^. Enrichment is plotted by zScore (y-axis) and significance (-log10(FDR q), color). Circle size denotes -log10(FDR q) value and color denotes enrichment significance (FDR q <0.05). **(G)** Lollipop plot for Fisher’s exact testing to evaluate overlap of DEGs and risk genes from multiple independently defined gene lists: Autism - SFARI gene database score 1 and 2 genes ^56^, Autism - Fu et al. ^57^, Autism - Satterstrom et al.^58^ as well as the collated autism risk genes.

We tested for transcriptomic overlap between datasets using two complementary analytic strategies: first, testing for overlaps among statistically significant differentially expressed genes (DEGs) in each condition; and second applying regularized canonical correlation analysis (rCCA) ^52^ as an efficient multivariate framework to simultaneously integrate threshold-independent gene-level logFC estimates between conditions across multiple cell types. This dual approach retained the distinctive strengths of both threshold-dependent and threshold-independent views of differential gene expression and helped identify the most robust results that jointly influence sex and autism or schizophrenia.

### Sex-biased genes in the cerebral cortex show a stronger overlap with differentially expressed genes in autism than in schizophrenia and highlight synaptic pathways enriched for autism risk genes

To provide an initial overview of transcriptomic overlaps we combined cell type DEGs within each dataset to define union lists of all unique genes (“global DEGs”) showing significant expression differences as a function of sex (N=1188), autism (N=5037), or schizophrenia (N=2455). In validation of these global DEG sets from snRNAseq data, they each significantly overlapped with their corresponding bulk RNAseq DEG sets from large independent studies (Sex Bulk ^53^-Pseudobulk ^45^ odds ratio (OR) = 1.64, FDR q = 2.48e-04, Autism Bulk ^54^ – Pseudobulk ^46^ OR = 1.78, FDR q = 4.04e-46, Schizophrenia Bulk ^55^ – Pseudobulk ^47^ OR =1.27, FDR q =6.76e-06) (**Figure S1A**). Our global snRNAseq DEG sets for autism and schizophrenia also recovered a well-established transcriptomic overlap between autism and schizophrenia (n = 1930, OR = 8.08, FDR q = 7.53e-70) as previously reported by bulk RNAseq ^48,49^ (**Table S2**).

Global DEGs from sex-differences snRNAseq (sex DEGs) showed a statistically significant overlap with those in autism (n = 599 shared genes, OR = 3.08, FDR q = 3.01e-70). This was significantly stronger (z = 4.22, p = 2.45e-05) than the significant overlap found from schizophrenia DEGs (n = 324 genes, OR = 2.04, FDR q= 2.49e-21, **Fig 1B-C**). Most of the sex DEGs that overlapped with autism and schizophrenia DEGs were autosomal (**Fig 1D**, 94.32% sex-autism, 96.30% sex-schizophrenia). The overlapping DEGs between sex and autism were relatively enriched for sex-linked X- and Y-chromosome genes (OR = 1.76, FDR q = 0.02). Six of these 31 sex-chromosome overlapping DEGs (19%) were autism risk genes — *NLGN4X*, *KDM6A*, *IL1RAPL2*, *CDKL5*, *USP9Y*, *NLGN4Y—* as defined by a collated set across multiple studies (**Methods**) ^56–58^. By contrast, the overlapping sex-schizophrenia DEGs were not enriched for sex-chromosome genes (OR=1.26, FDR q = 0.41; **Fig. 1E**). Importantly, the pattern of significant global DEG overlaps between sex and both neurodevelopmental conditions persisted when restricted to autosomal genes **(Fig S1B**-**C**). We also confirmed that the autism-sex-biased DEG overlaps observed in our primary analyses (**Fig 1A**) were robust to sampling of autism effects on cell type gene expression outside the frontal cortex ^54^ (**Fig S1D)**.

Overlapping DEGs between sex and autism were enriched for multiple terms, including regulation of synaptic structure or activity (zScore= 8.39, FDR q =4.02e-08), of synapse organization (zScore =8.28, FDR q =4.02e-08), and of axonogenesis (zScore= 7.19, FDR q = 1.27e-05; **Fig 1F**). The overlapping DEGs between sex and schizophrenia and between autism and schizophrenia were also enriched for multiple synaptic annotations (**Fig S1E-F**). These overlapping DEG sets also showed significant enrichment for autism risk. Specifically, the collated autism risk gene set was significantly enriched within the shared DEGs between sex and autism (OR = 2.63, FDR q = 6.23e-12) as well as the shared DEGs between autism and schizophrenia (OR = 3.03, FDR q = 2.44e-44) (**Fig 1G; Fig S1G**). This enrichment signal held for the individually curated sets of autism risk genes contributing to the collated set. We did not observe enrichment patterns for the collated set of schizophrenia risk genes in the shared DEGs between sex and autism (OR = 1.58, FDR q = 0.29) or its constituent sets, while schizophrenia risk genes were enriched in the overlapping DEGs between schizophrenia and autism (OR = 3.34; FDR q = 5.89e-07) (**Fig. S1H**). Together, these enrichment signals suggest that shared transcriptomic effects of sex and autism, sex and schizophrenia, and autism and schizophrenia converge on the synapse, confirmed by enrichment testing of global DEGs shared across all three datasets (**Fig. S1I**), and aspects of synaptic biology that are also enriched for autism risk genes ^59–61^.

### The overlap of differentially expressed genes in sex and autism varies by cell class and the cortical region where sex effects are quantified

Having defined a significant overlap between global DEG sets in sex and autism, we next asked if this global DEG set overlap was driven by specific cell classes in the autism dataset and specific region-by-cell class combinations in the sex dataset. To achieve this, we first used established rules ^64^ to harmonize cell type labels within the sex and autism datasets into six higher-order, shared cell classes for which DEG overlaps were determined: excitatory neurons (ExcNeu), inhibitory neurons (InhNeu), astrocytes (Astro), oligodendrocytes and oligodendrocyte precursor cells (Oligo/OPCs), microglia (Micro), and neurovascular cells (NeuVasc) (**Fig 2A**).

**Fig. 2:**
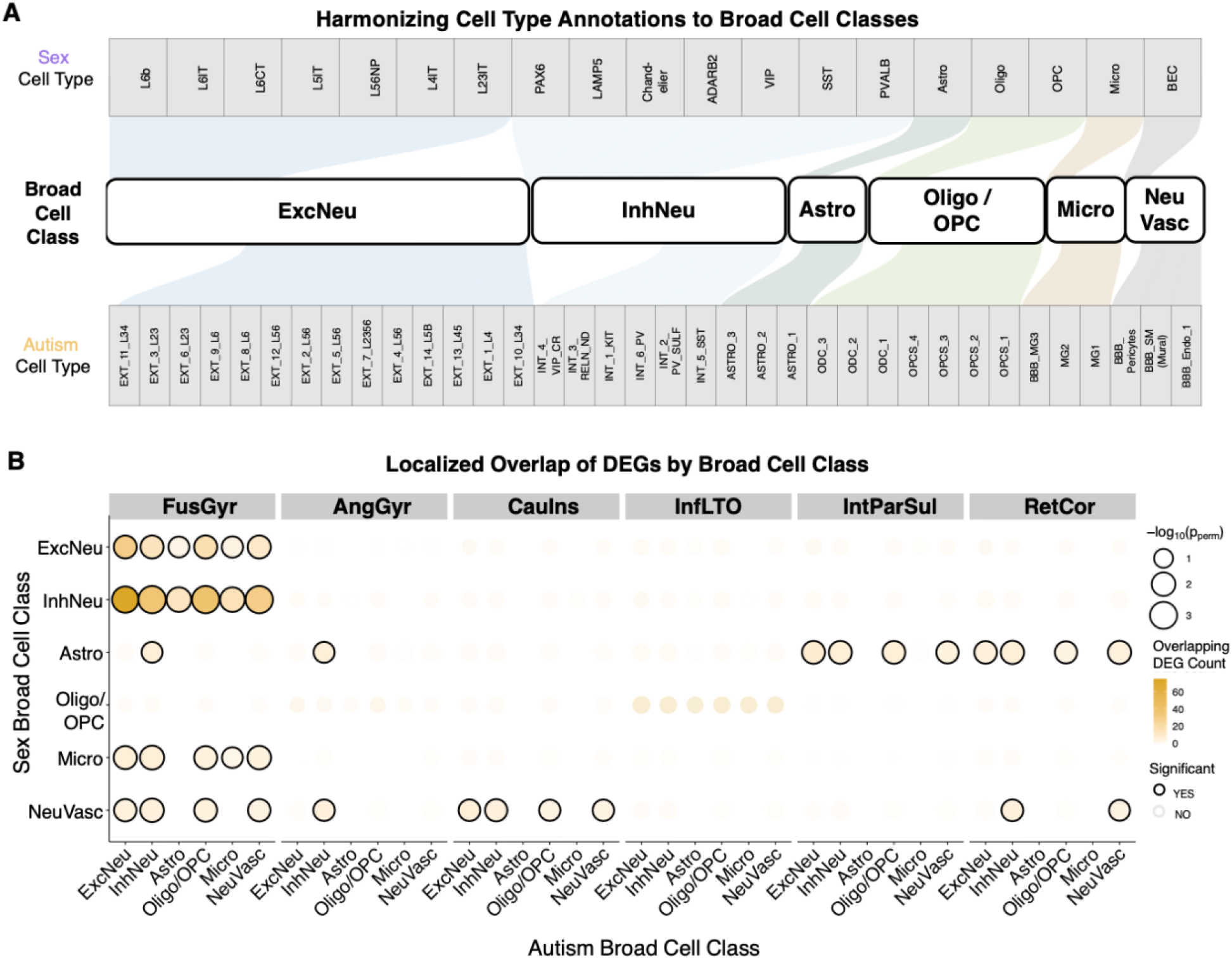
Overlap in differentially expressed genes between sex and autism highlights sex-biased patterns in FusGyr neurons. **(A)** Alignment of cell type annotations from sex ^45^ and autism ^46^ into broader cell classes. **(B)** DEG overlap by broad cell class based on intersection of DEGs. Empirical p-value derived by comparing the number of overlapping DEGs for each broad cell class comparison against the 1000 permutations of sex.

We quantified overlaps between cell class autism DEGs in frontal cortex and cell class sex DEGs in each of the six available cortical regions. These overlaps were tested for statistical significance against a null distribution of 1000 overlaps counts for the same region-by-cell class combinations from permutation of sex status before DEG calculation in the sex differences snRNAseq dataset (**Methods**). This procedure localized the significant global DEG set overlap between sex and autism (**Fig 1B-C**) to specific region-by-cell class combinations (**Fig 2B; Table S2**). The most statistically significant overlaps were seen between autism DEGs for multiple cell classes, and sex DEGs in the FusGyr neurons — especially sex DEGs in InhNeu. These overlaps remained statistically significant in sensitivity analyses considering only autosomal genes (**Fig S2A**). We also observed smaller — yet still statistically significant — overlaps between autism DEGs in ExcNeu, InhNeu, Oligo/OPCs and NeuVasc cells and sex DEGs in FusGyr Micro and NeuVasc as well as in AngGyr and CauIns Astro - although these weaker overlaps were more sensitive to exclusion of sex chromosome genes from analysis (**Fig S2A**). Within the significant localized DEG overlaps, we next characterized how direction of sex-bias (male-vs. female-biased) related to the direction of dysregulation in autism (up-vs. downregulated). Among DEGs overlapping between FusGyr InhNeu and autism ExcNeu, the direction of sex-bias was significantly associated with the direction of autism dysregulation (FDR q = 2.80e-2), with the largest number of shared DEGs for this comparison being those male-biased and upregulated in autism, followed by those female-biased and downregulated in autism, indicating an autism>control and male>female pattern of intersection (**Table S2**; **Fig S2B**). These findings held when analyses were restricted to autosomal genes (**Fig S2C**).

### Sex and autism share a multivariate axis of transcriptional effects that highlights specific cell classes and biological pathways and is enriched for autism risk genes

To complement the DEG-based analyses, which are sensitive to thresholding by arbitrary statistical cut offs for DEG definition and insensitive to the full multivariate pattern of logFC in each dataset snRNAseq database, we next integrated gene expression effects in sex and autism using a regularized Canonical Correlation Analysis (rCCA) ^65^ (**Fig 3A**). Briefly, we used rCCA to take the full gene-by-celltype logFC matrices created by differential gene expression analysis of snRNAseq in two datasets and define an ordered set of maximally correlated canonical variates between the two input datasets. Each canonical variate is the linear combination of logFC values in one dataset that is maximally correlated with its corresponding canonical variate in the other. We utilized a regularization approach (parameters lambda A = 0.1, B=0.1; **Fig 3B**) to stabilize the column loading. Regularization explicitly engages with the known correlational structure of logFC values between different cell types and region-by-cell type combinations for each dataset (**Fig. S3A-B**) ^66^. The rCCA framework produces multiple interpretable outputs, including correlation magnitudes and gene scores for each pair of canonical variates (A_n_, B_n_, **Fig 3B**). Use of rCCA to find aligned transcriptional effects of sex and autism not only avoids arbitrary binarization of logFC values (as is required for DEG overlap analysis), but is compatible with powerful rank-based methods for biological annotation, which have several advantages over testing for enrichments in variably sized sets of overlapping DEGs ^67^. Given that inclusion of sex chromosomes induces a major shift in logFC correlations within the sex difference dataset (reflecting disparate X- and Y-chromosome dosage between males and females), but not the autism dataset (**Fig S3C**) — we ran all rCCA analyses with and without inclusion of sex chromosome genes (**Fig S4, Methods**).

**Fig. 3:**
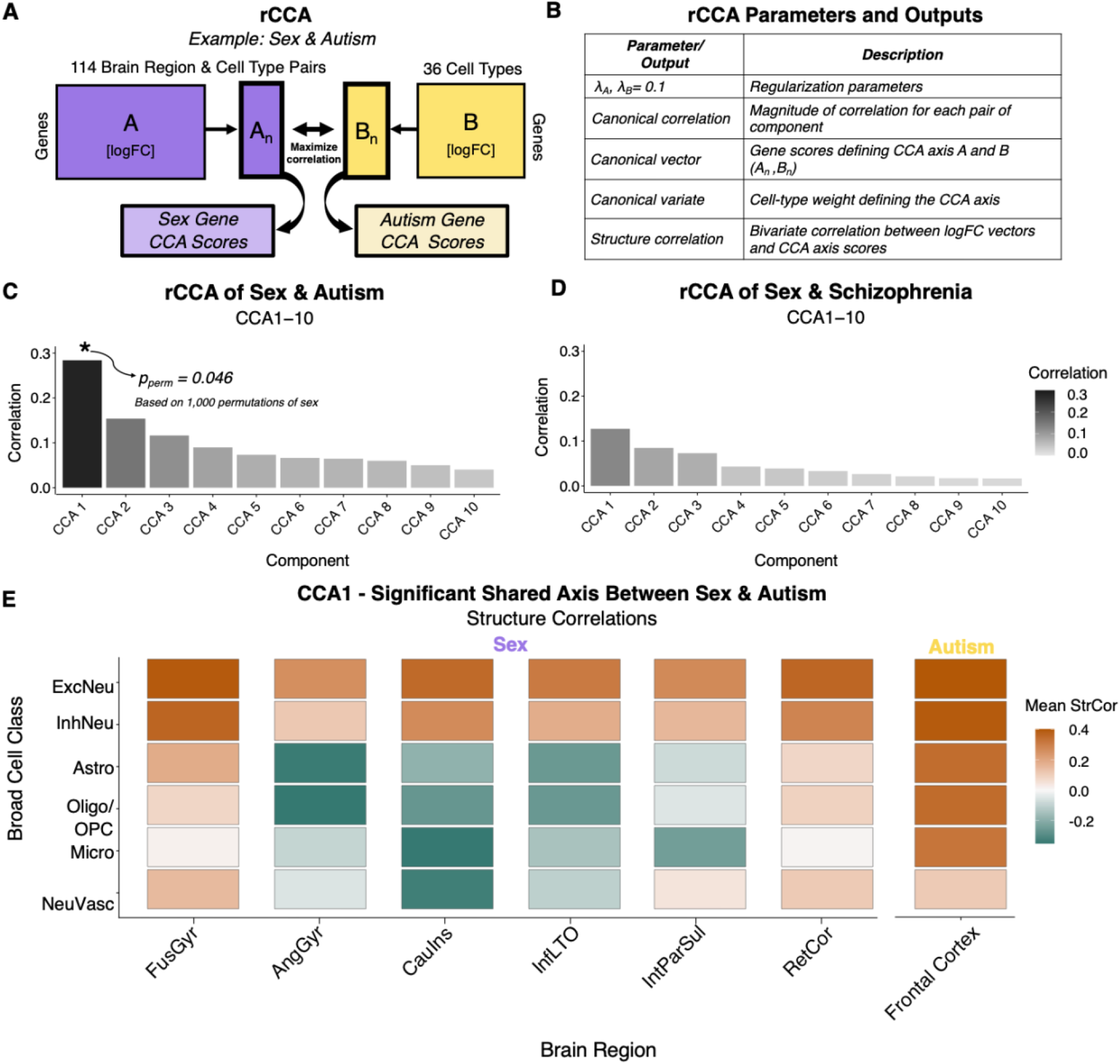
Regularized CCA reveals a statistically significant shared transcriptional axis between sex and autism, but not between sex and schizophrenia. **(A)** Schematic of rCCA, which is performed using matrices of logFC values for sex and each condition (autism and schizophrenia). Each matrix consisted of vectors of logFC values for genes across cell types (and brain regions for sex), with genes matched across datasets by gene name. Only genes present in at least one cell type were retained. Missing values for genes not detected in specific cell types were coded as NA. Regularized CCA was implemented using the CCA package ^65^ (v.1.2.2) to identify components capturing maximal correlation across datasets. CCA1 represents the pair of linear combinations (one per dataset) with the highest shared variance. **(B)** Outline of key CCA parameters and outputs. **(C, D)** Canonical components 1–10 for the sex-autism and sex-schizophrenia comparisons, respectively. CCA1 in the sex-autism analysis remained significant (p_perm_ <0.05) after 1,000 permutations of sex. **(E)** Mean structure correlations (Pearson correlation between logFC and CCA1 scores) of sex and autism (A_1_ and B_1_ respectively) per broad cell class.

Across all genes (n=16515), rCCA revealed a statistically significant CCA1 component between the sex and autism datasets (CCA1_(Sex, Autism)_ r= 0.28, empirical p_perm_ = 0.046 vs. 1000 permutations of the sex dataset, **Methods, Fig 3C**). This was significantly greater than the sex and schizophrenia CCA1 (z-score = 5.02, p = 5.18e-07; **Methods**), which did not reveal a significant shared axis (CCA1_(Sex, Schizophrenia)_ = 0.12, p_perm_=0.618, **Fig 3D**). The effect size between sex and autism was preserved in rCCA analyses run after exclusion of sex-chromosome genes: (CCA1 Auto _(Sex, Autism)_ = 0.27; albeit with a slightly raised permutation-based p-value of p_perm_ = 0.10, **Fig S4A**) and remained larger than that of the sex-schizophrenia CCA1 (CCA1 Auto _(Sex, Schizophrenia)_ = 0.12, **Fig S4B**). Sensitivity analyses supported the robustness of the sex-autism CCA1. Restricting to genes with complete case rows (n=3749) yielded a consistent signature (CCA1 Complete Case _(Sex, Autism)_ = 0.36; **Fig S4C; Table S3**), with highly correlated gene scores for sex and autism to the original CCA1 (mean r_CCA1-CCA1complete_ =0.92). Additionally, when sex– autism matrices were randomly downsampled 1000 times to match the dimensions of the sex– schizophrenia analysis (11145 rows × 25 columns), the sex–autism CCA1 never fell below the sex–schizophrenia CCA1 of 0.12 (**Fig S4D**).

Analysis of structure correlations of autism CCA1 gene scores within each cell class (mean gene score structure correlation with all constituent cell types, **Methods, Fig 3E**) revealed a broadly positive correlation of autism CCA1 scores with logFC values in autism vs. controls across multiple cell types (i.e. genes with low autism CCA1 scores tend to be downregulated in autism vs. controls across frontal cortex cell classes). For sex however, there were region and cell class specific correlations between sex CCA1 scores and logFC values in males vs. females (meaning that the direction of transcriptional alignment between sex and autism varies depending on cell class and region where sex effects are measured). Thus, genes at the negative CCA1 pole — which tend to be downregulated in autism across frontal cortex cell classes — tend to be female biased in FusGyr neurons, and male biased in glial cells across the AngGyr, CauIns and InfLTO junction (**Fig 3E**). A mirrored profile is seen for genes at the opposite positive pole of the shared transcriptional axis between sex and autism: these genes tend to be upregulated across cell classes in autism, male biased in FusGyr neurons, and female biased in multi-regional glial cell classes (**Fig 3E**). These structure correlations held on CCA1 analyses after exclusion of sex chromosome genes (**Fig S4E**).

We next focused on characterizing the significant CCA1 axis of shared transcriptional effects between sex and autism (**Fig 4A; Table S3**). A prominent feature of this axis was the high positive-ranking of Y-chromosome genes (CCA1 sex: normalized enrichment score (NES) = 2.18, FDR q = 8.17e-5 / CCA1 autism: NES = 1.84, FDR q = 0.02; **Table S4**). A similar pattern was seen for gene scores on the weaker and statistically insignificant CCA1 from comparison of gene expression effects of sex and schizophrenia (CCA1 sex: NES = 2.46, FDR q = 1.58e-29 / CCA1 schizophrenia: NES = 1.87, FDR q = 0.11; **Fig S5A; Table S4)**. Indeed, Y-linked genes received identically ranked mean CCA1 scores for the sex-autism and sex-schizophrenia analyses (**Fig S5B**). By contrast, X-linked genes did not show a significant enrichment amongst genes ranked by CCA1 score for the sex-autism analysis (CCA1 sex: NES =1.14, FDR q = 0.48 / CCA1 autism: NES = 1.13, FDR q = 0.25) (**Table S4**). Profiling Y-linked gene expression in all datasets revealed the expectedly ubiquitous and large difference in Y-linked gene expression in the contrast between males and females (**Fig S5C**), alongside small, sparse and directionally variable differential expression in the case-control contrasts for autism and schizophrenia (**Fig S5D,E**). These observations indicate that the high-ranking of Y-linked genes on both the sex-autism and sex-schizophrenia CCA1 components analyses reflects the aliasing of sex with Y-chromosome dosage rather than a substantial patterning of altered Y gene expression in autism or schizophrenia that contributed to their shared transcriptional axis with sex. Supporting this conclusion, sex-autism CCA1 gene scores were highly correlated between rCCA analyses run before and after exclusion of sex chromosome genes from analysis (mean r = 0.94, **Fig S4F,G**).

**Fig. 4:**
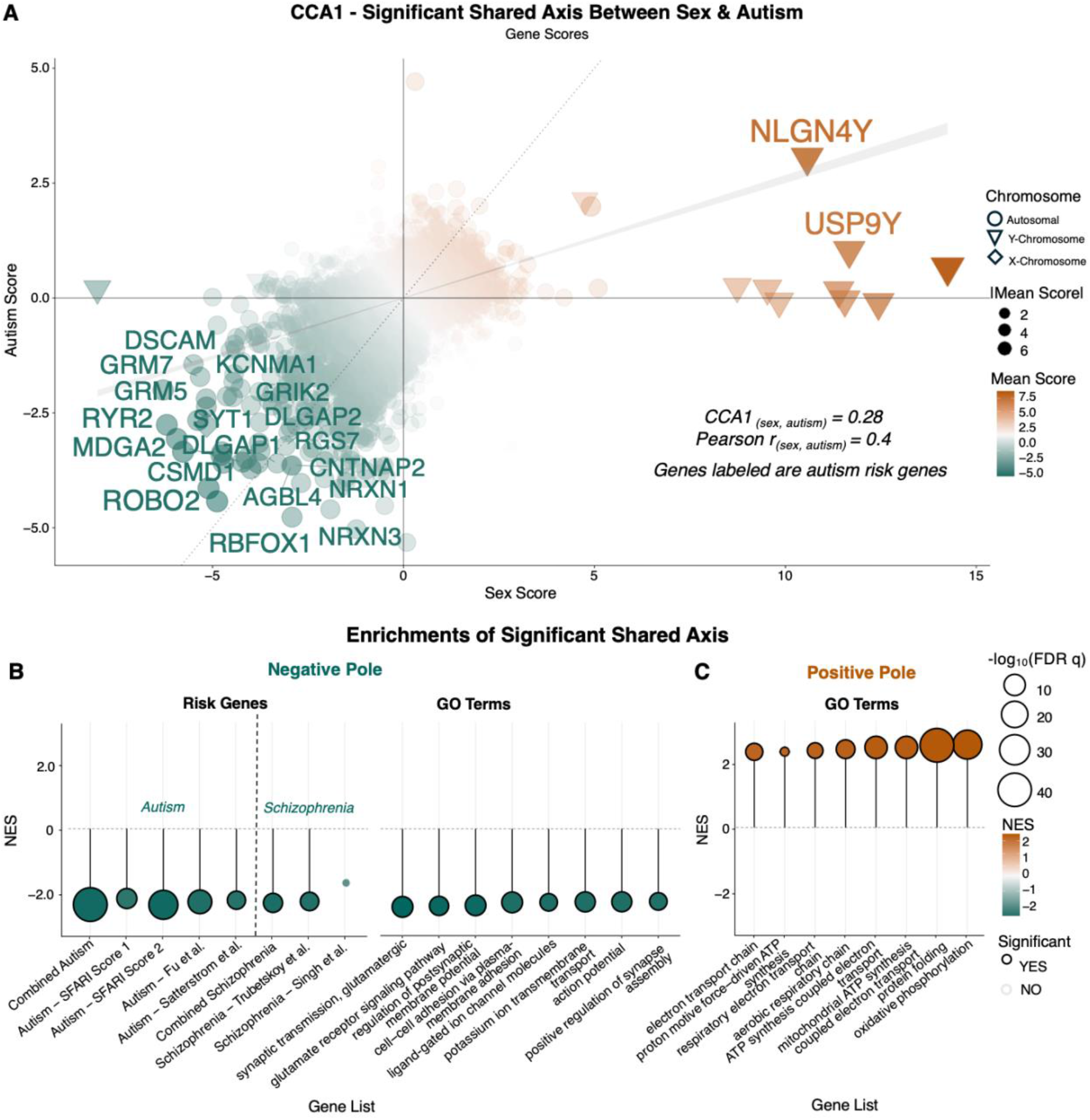
Properties of the shared CCA1 transcriptional axis between sex and autism. **(A)** Scatter plot of CCA1 gene scores for sex (x-axis) and autism (y-axis). Each point represents a single gene; shape encodes chromosomal origins: autosomal (circular), X-chromosome (diamond), Y-chromosome (triangle). Top 20 autism risk genes (from collated autism risk gene set) were labeled; the top were defined by greatest absolute mean score. Color encodes mean score with high positive CCA1 score (tan) and low negative CCA1 score (teal). The solid grey line indicates the line of best fit and the dotted grey line indicates slope of 1. **(B)** Lollipop plot for Enrichment analysis of neurodevelopmental condition risk genes and Gene Ontology (GO)^62^ Biological Process (BP) terms. GO terms were selected for the positive and negative pole of the results by normalized enrichment score (NES) of significant (FDR q <0.05) values.

We next biologically characterized the stable and statistically significant shared CCA1 axis of transcriptional change between sex and autism through functional enrichment testing. To simplify rank-based annotation of this CCA component by gene set enrichment analysis (GSEA)^68^; **Methods**), we derived a single gene ranking for annotation by averaging the sex and autism CCA1 scores into a single number (“CCA1 score”) for each gene. The CCA1 scores defined a shared axis that was biologically distinct at both poles. GSEA identified more significant enrichments at the negative vs. positive CCA1 pole **(Fig S6A, B; Table S4)**. Genes at the negative sex-autism CCA1 pole were significantly enriched for multiple glutamatergic synaptic signaling annotations and risk genes for both autism and schizophrenia (although more statistically-significantly so for autism vs. schizophrenia risk genes; **Fig 4B, Table S4**). We found the neuronal autism risk genes had significantly more negative CCA1 scores than gene expression regulation autism risk genes (Wilcoxon rank-sum test: W = 1039; p = 1.19e-04), as annotated by Satterstrom et al. ^58^, revealing that autism risk genes related to the synapse annotated were preferentially associated with the negative sex-autism CCA1 pole. In contrast, genes at the positive pole were significantly enriched for mitochondrial energy metabolism (**Fig 4C**) and immune annotations (**Fig S6E**) but lacked any enrichment for risk genes (**Table S4**). Both the negative and positive poles were enriched for sex-autism shared global DEGs (negative: OR = 9.46, p = 4.27e-10; positive: OR = 5.04, p = 1.94e-04), indicating that the threshold-dependent and threshold-independent analyses converge. We next tested enrichment of the localized DEGs from the sex-DEGs in the fusiform gyrus inhibitory neurons and autism-DEGs in the excitatory neurons. The negative pole showed a trend towards enrichment of these shared DEGs (OR = 5.84, p =0.05), whereas the positive pole showed a significant enrichment (OR = 22.30, p = 7.99e-07). This agrees with the DEG-based localization, in which most of these shared DEGs were male-biased and upregulated in autism (**Table S2, Fig. S2B**), consistent with the positive pole’s stronger enrichment.

Considered alongside structure correlations for the sex-autism CCA1 gene scores (**Fig 3E**), these findings indicate that genes at the negative sex-autism CCA1 pole — which are enriched for autism genetic risk and glutamatergic synapse annotations— tend to be downregulated across almost all cortical cell classes in autism, female biased in multiple FusGyr cell classes, and male biased in multiple glial cell classes across the AngGyr, CauIns and InfLTO junction (**Fig S7A-B**). Correspondingly, genes at the positive sex-autism CCA1 pole— which lacked enrichment for autism genetic risk but tagged energy metabolism and immune terms— showed the opposite expression profile: upregulated in autism across all cell classes, male biased in multiple FusGyr cell classes and female biased in almost all glial cells across the AngGyr, CauIns and InfLTO junction (**Fig S7A-B**). Thus, sex and autism converge on a shared transcriptional axis with two opposing poles, the negative pole defined by autism risk and synaptic signaling, downregulated in autism, female biased in FusGyr, and male biased across other cortical brain regions; and the positive pole defined by metabolic and immune function, upregulated in autism with the reciprocal sex-bias pattern.

### Molecular networks at the transcriptional overlap between sex and autism

We next harnessed protein-protein interaction (PPI) analysis to test for biological interactions amongst genes at the negative pole of the sex-autism CCA1 axis (bottom 0.5% CCA1 gene score), which showed enrichment for autism risk genes (**Fig 5**). Parallel results are provided throughout for the positive CCA1 pole in supplementary material (**Fig S8**).

**Fig 5:**
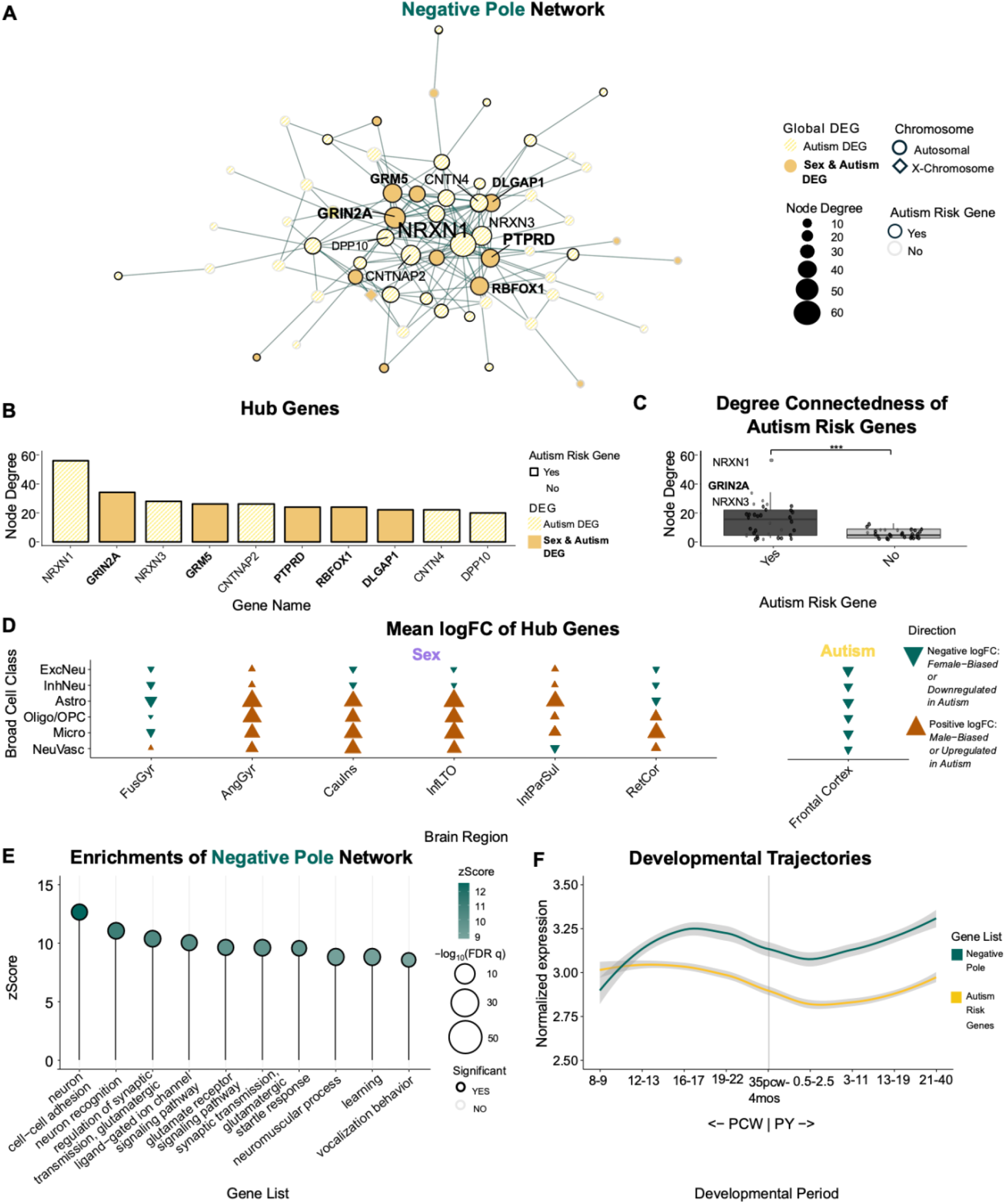
Biological characterization of the shared transcriptional effects of sex and autism that are enriched for autism risk genes (CCA1 negative pole). **(A)** Protein-protein interaction (PPI) networks for the top 0.5% of the negative pole of CCA1 mean scores. Nodes are categorized by chromosomal origin (autosomal: circle; Y-linked: triangle; X-linked: diamond) and outlined by SFARI score (score 1: black; score 2: dark grey; other/none: light grey/white). Hub genes are labeled, with bold labels indicating genes that are DEGs in both sex and autism. **(B)** Bar chart of node degree for the ten most connected genes (hubs). Fill indicates DEG status (hatched: autism DEG; solid: sex and autism DEG), and outlines indicate autism risk gene status. **(C)** Boxplot comparing the distribution of nodal connectedness for genes, grouped by whether each gene is included in the collated autism risk gene list. Wilcoxon rank-sum test was used to compare median connectedness between the two groups; p-values are reported, with p < 0.001 denoted as ***. **(D)** Dot plot of the mean logFC of hub genes to project the CCA1 directions back to the gene expression changes observed for sex and autism. Color and shape encode the direction of the mean logFC (across hub genes and fine cell types within the broad categorization) showing positive logFC - male-biased or upregulated in autism (upward facing triangle; dark tan) and negative logFC - female-biased or downregulated in autism (downward facing triangle; dark teal), showing female-biasing of negative network hub genes for FusGyr neurons and male-biasing for AngGyr glial cells, consistent with structure correlations. **(E)** Lollipop plot for functional enrichments of GO Biological process ontologies from the negative pole compared to the CCA1 background using clusterProfiler ^63^. **(F)** Developmental trajectory curves of cortical normalized gene expression for negative pole and for collated autism risk genes across nine developmental period from the PsychENCODE database^70^ from post-conceptional week (PCW) 8 to year 40. The shaded region indicates 95% confidence intervals.

Genes at the negative pole of the sex-autism CCA1 (N=79) were enriched in PPI edges compared to a degree-matched null permutation (**Methods**, p_perm_ < 0.001). Of these, 62 (78.5%) of these mapped genes coalesced into a single connected network comprising 159 interactions (**Fig 5A**). Network analysis revealed that the ten most connected nodes (“hubs”) were *NRXN1*, *GRIN2A*, *NRXN3*, *GRM5*, *CNTNAP2*, *PTPRD*, *RBFOX1*, *DLGAP1*, *CNTN4*, and *DPP10* (**Fig 5B**). All ten hub genes were differentially expressed in autism and half were differentially expressed in sex (**Fig 5A-B**), linking the CCA-based, threshold-independent analysis with the DEG-overlap, threshold-dependent analysis. The network was strongly enriched for autism risk genes (34 of 62 genes, 54.8%, versus 7% of the CCA1 background; OR = 16.2, p < 0.001), and each of the ten network hub genes were risk genes. This indicates that the convergent signal between sex and autism captured by the negative pole concentrated on highly connected, established risk genes. This reflected a broader tendency within this network that autism risk genes were more interconnected than chance, based on retesting connectedness with shuffled autism risk gene assignment (**Methods**) (p_perm_ <0.001 in each degree bin from two-to-ten), and showed significantly higher node degrees than non-risk genes (Wilcoxon rank-sum test: W = 718, p = 5.27e-04; **Fig. 5C**).

In line with expectations from rank-based enrichment analyses of sex-autism CCA gene scores (**Fig 4B; Fig S7**; **Table S4**), negative-network hub genes were downregulated in autism, female biased in FusGyr neurons, and male biased across cell classes in AngGyr (**Fig 5D**). These genes were also strongly enriched for neuronal communication terms, including neuron cell-cell adhesion (FDR q = 2.49e-04), neuron recognition (FDR q = 3.41e-05) as well as other domains, including learning (FDR q = 3.48e-05), vocalization behavior (FDR q= 6.64e-03), and startle response (FDR q = 1.39e-03) (**Fig 5E)**.

Given the important developmental dynamisms of sex and autism effects, i.e. pre-natal expression of autism risk genes, we predicted genes at the negative pole would show peaks of expression during specific developmental windows, particularly pre- and early postnatal period and puberty. Genes in the negative pole showed dynamic expression patterns similar to the trajectories of the collated autism risk genes: increasing throughout prenatal development, with a slight reduction postnatally and a slight rise in late adulthood (**Fig 5F**).

Finally — noting this prenatal dynamism in expression of genes defined by our analysis of shared transcriptional effects of sex and autism in adulthood — we asked how the signal of sex effects on cell type gene expression in adulthood related to those described for the prenatal and postnatal human brain ^69^ (**Methods**). Specifically, we compared the ranked sex differences from our dataset from each region-by-cell class by logFC from our analysis with the DEGs by lineage and sex from the developmental dataset starting at the second trimester of the prenatal period. These analyses revealed that sex-biased expression in the adult fusiform gyrus — which shows particularly strong overlaps with autism gene expression changes — is more concordant with sex effects on developmental brain gene expression than other regions in our dataset (**Fig. S9**). This suggests that the fusiform gyrus’s alignment with female protective effects from adulthood sex differences may partly reflect stable sex differences across other developmental windows, which is important as autism risk genes and broader gene expression differences may exert particularly strong influences during early developmental windows.

## Discussion

Our study integrates newly published single nucleus RNAseq studies of human cortex to reveal transcriptional overlaps between sex and autism, which localize to specific brain regions, cell classes and molecular pathways, and show enrichment for known neurodevelopmental risk genes. Our findings advance the understanding of biological intersections between sex and autism in three main directions as detailed below.

First, by combining differentially expressed genes across cell types, we show that these new snRNAseq results complement several findings from prior bulk RNAseq studies of transcriptomic overlaps between sex and autism ^23,38,44^. Specifically, previous reports, primarily from bulk cortical RNAseq studies have reported that: female-biased genes are enriched among synaptic genes downregulated in autism and among neuronal/synaptic genes; and male-biased genes are found to be enriched among genes upregulated in autism and among microglial/immune genes ^38,44,71,72^. Our results corroborate and extend these observations with cell type resolution for synaptic pathways. We confirm that synaptic genes are indeed enriched amongst those that are both female-biased and down-regulated in autism within neurons. However, these same synaptic pathways are enriched amongst genes that are male-biased within glial cell types across multiple brain regions. In glia, these pathways are thought to support glial-neuronal interactions at the “tripartite synapse” ^73^. Speculatively, if female-biased expression of these genes in neurons imparts resilience to genetic risk for autism, and male-biased expression of these genes in glial cells reflects a male-biased criticality of function, then this could set up two compounding pathways for sex-biased resiliency to genetic risk for autism at the synapse. We also recover the signal from prior bulk RNAseq studies of converging transcriptional signatures between sex and autism in immune-related biology ^44,74,75^. However, these enrichments were comparatively weaker in snRNAseq data, and no immune GO terms were enriched in the shared DEGs between sex and autism, which may relate to the theorized non-genetic etiology of immune-gene dysregulation ^76^, or subtle cell type proportion effects of sex and autism biasing bulk-RNAseq findings. Notably, we find that the nature of this overlap between sex and autism effects on immune-gene sets also differed by cellular context, a finding that would have been masked in bulk tissue. We show that immune pathways tend to be upregulated in autism broadly across all cell classes, upregulated in males within fusiform gyrus and caudal insula neurons, but upregulated in females within glial cells across many brain regions. This observation for immune pathways also invites the idea that intersections of sex and autism could potentially involve convergences of different mechanisms between interacting cell types. Taken together, these findings from new snRNAseq datasets further strengthen the idea that there is significant biological intersection between sex and autism at the level of gene expression. Strikingly, both complementary analytic approaches in our paper — the DEG-based and the CCA analysis — find that the strength of this intersection appears to be more prominent than that between sex and schizophrenia, a difference which aligns with the greater sex bias in autism vs. schizophrenia prevalence ^4,51^.

Second, our use of snRNAseq measures of sex effects on gene expression from multiple brain regions lets us test if cell type transcriptional alignments between sex and frontal cortex signatures of autism vary by the brain region where sex effects are estimated. This test relies on measures of autism effects in frontal cortex alone because the field currently lacks multiregion snRNAseq datasets in autism. However, large-scale multiregion studies using bulk RNAseq suggest that transcriptomic signatures of autism are highly correlated across cortical regions^54^.We find that transcriptional effects of autism as captured by snRNAseq data in the frontal cortex show regionally-varying alignment with sex effects on cortical gene expression — with the strongest transcriptomic overlap being observed with sex effects on gene expression in the fusiform gyrus. This alignment - which echoes the prominently large and distinctive nature of sex effects on cell type gene expression in the fusiform gyrus as compared to the other regions under analysis ^45^ - is particularly striking given the fusiform’s well-established role in facial perception ^77,78^, a domain which has been implicated in both sex differences ^79,80^ and autism ^81,82^. Further, the fusiform gyrus lies downstream of occipital visual cortex within the ventral visual stream, which may be meaningful as the visual network has been demonstrated to show the greatest transcriptomic disruption in autism ^54^. The region and cell class dependence of transcriptional overlaps between sex and frontal cortex signatures of autism may help to reconcile some of the conflicting reports from prior studies ^38,83^. For example, other studies have reported female-biased (rather than male-biased) expression of immune genes and male-biased (rather than female-biased) expression of synaptic genes ^75,84^, with no substantial correlation to autism signatures in that context ^85^. The directional variability we observe from different brain regions and cell types in sex may help explain why prior reports of sex effects on autism risk have been heterogeneous, which underlines the need for greater spatial sampling of brain tissue in future work. Additionally, only a subset of localized overlaps based on DEGs involved the same cell types in sex and autism, a finding that also would have been masked in bulk tissue, and interpretation of this differs based on what the cell types are. By pursuing a more comprehensive sampling of brain regions and cell types, future work will be able to further probe the model evoked by our current findings: connections between the neurobiology of sex and autism may be more prominent, and perhaps mechanistically pertinent, in some brain regions and cell types than others.

Third, we use our region and cell-class resolved view of transcriptional overlaps between sex and autism as input to protein-protein interaction (PPI) networks — revealing specific sets of interacting partner molecules that are not just enriched for convergent transcriptomic effects of sex and autism, but also for autism risk genes. We resolve a synaptic PPI network — which tends to be female biased in neurons and male biased in glial cells — that is highly enriched for genes that increase autism risk when haploinsufficient ^86,87^. Interestingly, while autism risk genes have been characterized into multiple domains, e.g. chromatin regulation, and synaptic ^58,88^, the overlapping effects of sex and autism concentrated on synapse-related risk genes. Some of these autism risk genes have previously been shown to exhibit region-specific expression across the brain ^89,90^, and our evaluation further revealed that while autism risk genes are ubiquitously downregulated in autism across most frontal cortex cell types, their sex-biasing exhibits two modes, with patterns of female biasing, consistent with female-protective effects, present in fusiform gyrus and retrosplenial cortex, and patterns of male biasing distributed across angular gyrus, caudal insula, and inferior later temporal occipital junction. This raises the possibility that it is the female-biased expression of risk genes in certain brain regions that may be especially salient to the risks and biology of autism. Future work is required to systematically evaluate the regional expression patterns of these risk genes across development and to fully delineate the meaning of the direction of their sex biasing.

Our findings should be considered in light of several important caveats and limitations. First, we operationalize sex as a binary variable based on genomically confirmed sex chromosome dosage, but there are other criteria by which sex can be measured, and there are groups of individuals who have mixed profiles of these criteria — meaning that sex is not strictly binary ^91^. Second, we do not incorporate measures of gender — a complex multidimensional psychosocial construct that is entangled with sex and may account for some measured associations between sex and neurobiology. Third we characterize transcriptomic features of autism using associations between gene expression and a binary grouping of donors into those that met criteria for autism vs. those that did not. This approach is appropriate given the current sample sizes of postmortem brain snRNAseq data in autism but cannot engage with the well-established heterogeneity of autism at clinical, neurobiological and genetic levels of analysis ^92^. Additionally, the autism snRNAseq data used here was exclusively from the frontal cortex. Previous work in bulk tissue found that signatures of autism are correlated across the cortex ^54^, and we provide preliminary evidence that our findings hold using smaller non-frontal snRNAseq datasets in autism. However, it will be important for future work to further characterize overlaps of sex and autism across multiple brain regions. Fourth, expanding datasets will also make it possible to more fully probe how factors such as developmental stage and brain region might modulate cell type transcriptomic overlaps between sex and autism, especially regarding the more developmentally stable sex differences in fusiform gyrus compared to other brain regions. These expanded datasets would also benefit from investigating the sex-specific signatures of autism. Finally, our study of postmortem gene expression in humans is necessarily descriptive. While this design provides a powerful screen for potential transcriptomic overlaps between sex and autism, other study designs in model system will be needed to experimentally test the hypotheses raised by our work.

Notwithstanding these limitations, our work captures a significant intersection of gene expression for sex and autism and places this intersection in a more granular cellular context than was previously possible. We find overlaps between sex and autism effects within autism risk genes, which previous reports did not find ^38,93^. Our findings strengthen prior theories that synaptic and immune pathways may be points of biological convergence between sex and autism but show that the nature of this convergence — its magnitude and paired directions of fold change in sex and autism — varies as a function of both brain region and cell type.

## Methods

### Differential gene expression across datasets

Cell type resolved measures of differential gene expression in autism, and schizophrenia were drawn from recently-published landmark studies by Wamsley et al. ^46^ and Ruzicka et al ^47^, respectively. Both of these studies applied *limma* ^94^ to single-nucleus RNA-sequencing (RNAseq) data from samples of frontal cortex in sex-balanced case-control samples (autism cases = 33 [males = 29, females =4], autism controls = 30 [males =23,females =7]; schizophrenia cases = 65 [males = 42, females = 23], schizophrenia controls = 75 [males = 43, females =32]). Both studies used group contrast where cases were compared to controls, so that positive logFC indicates higher expression in autism or schizophrenia. We generated analogous measures of cell type differential expression between males and females by applying *limma-voom* ^94^ to the recently-published snRNAseq dataset from DeCasien et al ^45^ which spans 169 cortical samples from 15 males and 15 females across 6 cortical regions [FusGyr, InfLTO, CauIns, IntParSul, RetCor, and AngGyr]. Single-cell library preparation, read mapping, and quality-control was previously conducted on this dataset by DeCasien et al. ^45^. For differential gene expression, the counts were aggregated and normalized across cells for each cell type and sample to create a pseudobulked counts matrix. Differential gene expression for sex was run accounting for relevant covariates chosen by the original analysis ^45^, including post-mortem interval, age, RNA-integrity number, total number of reads, and total number of cells. Sex-associated differential expression was assessed using a contrast comparing male versus female samples, thus positive logFC indicates a male bias and negative logFC indicates a female bias.

### Differentially expressed gene overlaps

Differentially expressed genes (DEGs) were defined in the autism and schizophrenia datasets according to the published criteria applied to *limma-voom* outputs in the respective publications (False Discovery Rate (FDR) q < 0.05; |logFC| > 0.2 and >0.1 in the autism ^46^, and schizophrenia ^47^ datasets, respectively). A more relaxed statistical threshold was applied to the sex-differences dataset (FDR q < 0.1), in line with previous work ^95^, to ensure capture on expectedly small sex effects autosomal gene expression. Prior work in this dataset has established that FDR q < 0.05 thresholds to *limma-voom* output primarily detect sex DEGs on the dosage-disparate X- and Y-chromosome, and fail to recover those distributed sex effects on autosomal gene expression that are otherwise detectable by multivariate adaptive shrinkage (MASH) ^96^ methods ^45^.

Pairwise DEG overlaps were analyzed at two levels of resolution. First, DEGs were collapsed across all cell types (“global” DEGs) in each of the three datasets and used to test for enriched overlaps for each of three pairwise comparisons by Fisher’s exact test vs. the respective background: sex and autism, sex and schizophrenia, and autism and schizophrenia. To compare the odds ratio of the tests, we followed standard methodology for comparing two independent estimated quantities, dividing the difference between the log-transformed odds ratios by the standard error of that difference, calculated as the square root of the sum of the squared standard errors of each log odds ratio. The resulting z-statistic was compared to the standard normal distribution to obtain a two-sided p-value^97^.

Fisher’s exact testing p-values were adjusted for multiple comparisons via Benjamini-Hochberg FDR correction. Each global overlap set was then tested for functional enrichments with *enrichGO* from the *clusterProfiler* package ^63^ compared to their respective shared background, with minGSSize = 10 and maxGSSize = 500. Second, for the sex and autism comparison, DEGs overlaps were also assessed at finer resolution — for collapsed DEG sets within each broad cell class (“localized DEGs”). The localized overlapping DEG sets were evaluated by comparison to a null distribution of localized overlap counts using sex DEG sets from 1000 runs of *limma-voom* ^94^ on the sex differences snRNAseq dataset after sex permutation by donor. For all cell class combinations showing significant DEG overlaps (p_perm_ < 0.05), we quantified the proportion of overlapping DEGs within the 4 possible combinations of mean logFC direction (meanlogFC across the constituent cell types within each cell class) between datasets: (1) male>female and upregulated in autism; 2) male>female and downregulated in autism; 3) female>male and upregulated in autism; or 4) female>male and downregulated in autism. We then tested the enrichment of each of these four directional combination sets using Fisher’s exact test, with p-values adjusted for multiple comparisons via Benjamini-Hochberg FDR correction ^98^. All DEG overlap analyses were conducted with and without exclusion of sex chromosome genes as a sensitivity analysis.

### Regularized canonical correlation analysis

Regularized canonical correlation analysis (rCCA) was used to identify multivariate patterns of shared gene expression changes between datasets – thereby taking advantage of the full distribution of observed logFC values of all genes, without imposing arbitrary statistical thresholds for defining DEGs ^99,100^. This multivariate approach also directly accommodates and models correlated fold change profiles between cell types for each condition, e.g. as recently reported for sex effects on gene expression ^45^.

The inputs of rCCA were gene-by-cell type matrices of logFC values, with genes as rows and cell types as columns. We applied rCCA twice — once for the sex and autism comparison, and again for the sex and schizophrenia comparison. For the sex matrix (matrix A), columns represented each cortical region-by-cell type combination; for the autism and schizophrenia matrices (matrix B), columns represented cell types. Cell type labels were harmonized into broad classes across datasets, but fine-grained differential-expression contrasts were retained within each matrix. Genes were matched between dataset pairs by gene symbol; genes absent from either matrix were excluded, and missing gene-by-cell type estimates were treated as missing values. The sex–autism analysis included N = 16,515 matched genes.

The regularization aspect of rCCA prevents overfitting by penalizing the weights (λ_A_ and λ_B_) applied to columns for definition of canonical axes (Eq. 1; ^66^).

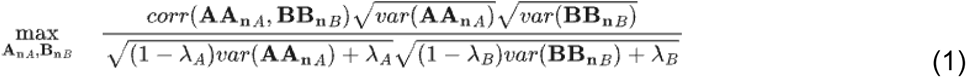

To select appropriate regularization parameters, we implemented a grid search optimization by calculating the mean correlations across canonical covariates across a range of regularization values from 0.1 to 1 in increments of 0.1 for both λ_A_ and λ_B_. The optimal parameters were defined as the values of λ_A_ and λ_B_ that yielded the maximal mean canonical correlation across dimensions. Analyses were performed using the *rcc* function from the CCA R package (v.1.2.2) ^65^. The outputs of rCCA included the vectors of gene scores for each canonical axis corresponding to each input matrix. To test for statistical significance of rCCA components, we compared observed magnitudes of correlation across the canonical axes to a distribution of null correlations generated after re-running rCCA 1000 times using sex logFC matrices from permutations of the sex variable.

To determine which cell types and region-by-cell type pairs contributed to each canonical axis, we calculated structure correlations. For each column of an input matrix, a structure correlation is computed as the Pearson correlation across genes between columnar logFC values and gene scores on the canonical variate ^66^. Structure correlations were averaged across the constituent fine cell types in each dataset (and nested by region for the sex dataset) to map how shared axes of gene expression change manifest across the cell classes and regions represented in our datasets.

To compare the magnitude of CCA1 of the different tests, we followed the same procedure for odds ratio comparison, but without log transformations ^97^. To determine confidence intervals for the CCA1 stability, we generated 1,000 bootstrap samples for the sex-autism and sex-schizophrenia analyses using row resampling. The outputs of rCCA, i.e. statistical significance of rCCA components, their structure correlations and gene scores, were also tested for stability after exclusion of sex chromosome genes from analyses.

### Enrichment testing

We ran Gene Set Enrichment Analysis (GSEA) using the fgsea (*v 1.36.2*) R package ^68^ for biological annotation of the statistically significant shared CCA1 axis between sex and autism. For these analyses, genes were ranked by the mean gene score of the two canonical variates. We tested for rank-based enrichment of the following gene sets: Gene Ontology ^62^ functional pathways, SFARI gene database ^56^ score 1 and score 2 genes, Fu et al. ^57^, Satterstrom et al. ^58^, Trubetskoy et al. ^101^ and Singh et al. ^102^, as well as the union of each condition’s individual risk gene sets, which generated a collated risk gene set for autism and schizophrenia. Statistical significance was determined using multilevel Monte Carlo (nperm =1000) and corrected for multiple comparisons via Benjamini-Hochberg FDR ^98^ correction across all tested gene sets.

### Protein-Protein Interactions

Protein-Protein Interaction (PPI) networks were determined using the STRING database with the STRINGdb (v.2.20.0) R package ^103^. PPI networks for the CCA1 scores were derived from the top 0.5% of the scores for the positive and negative pole (generating gene sets for PPI networks analysis). After removing unmapped entries, edges were defined as protein-protein interactions between mapped genes with a score threshold >400, following default parameters. The resulting networks were modeled using the igraph (v 2.3.3) ^104,105^ R package to calculate node degrees for each gene.

Network connectivity was defined as the number of edges within the induced subgraph of each gene set (n_edges). To test whether observed connectivity was significantly elevated relative to chance, we implemented a node degree-matched permutation procedure. First, we constructed a PPI network from the full CCA1 gene background (n = 16,515 genes). For every gene in this background network, we computed its degree (degree (graph_network, mode = "all")), and then all 16,515 genes were partitioned into 30 bins based on the quantiles of this degree distribution. We generated a null distribution by drawing 10,000 permuted gene sets matched to the degree profile of the genes from the respective network being tested. We extracted an induced subgraph from the full PPI network (induced_subgraph), and the same connectivity statistic (n_edges) was computed as for the observed gene set, which was then used to calculate an empirical p-value (p_perm_). To test whether autism risk genes were more interconnected than other genes in the same set, risk-gene labels were randomly reassigned among the network genes. Null sets of the same size as the observed autism risk-gene set, annotated based on the collated autism risk genes, were drawn from all network genes, matched on full-network degree using (2–10) quantile bins, with 10,000 permutations per setting. The statistic and P value were calculated as above. To assess sensitivity to binning, the analysis was repeated across 2 to 10 bins.

Genes that mapped onto the respective PPI network were submitted for enrichment testing of Gene Ontology^62^ terms using *enrichGO* from the clusterProfiler (v 4.18.4) R package ^63^ compared to the CCA1 background (n=16515 genes) with minGSSize = 10 and maxGSSize = 500. Lastly — for genes along the poles of sex-autism CCA1, we also profiled trajectories of developmental expression in bulk RNAseq using previously published methods Stauffer et al. 2023 ^106^ as applied to the PsychENCODE dataset of regional cortical gene expression spanning 8 post-conception weeks-40 years of age ^70^. For an input gene set of interest this methods maps its mean-centered cortical expression across 9 developmental windows (window 1: post-conceptional week (PCW) 8-9, window 2: PCW 12-13, window 3: PCW 16-18, windows 4: PCW 19-22, window 5: PCW 35-PY 0.3, window 6: post-natal year (PY) 0.5-2.6, window 7: PY 2.8-10.7, window 8: PY 13-19, window 9: PY 21-64).

Stability of sex differences from adulthood, as tested in DeCasien et al. ^45^, compared to developmental sex differences were evaluated by inputting the ranked adulthood sex differences logFCs by broad cell class and brain region into enrichment testing of DEGs from the developmental sex differences characterized by Velmeshev et al ^69^. This reference evaluates sex differences across developmental stages starting at the second trimester to determine sex effects on cellular lineage and development. We ran functional enrichment analysis with *fgsea* following the same procedure as described above. The gene lists for testing were the differentially expressed genes by both lineage and direction of sex-biasing. Enrichment values with positive indicates male biased in adulthood and negative indicates female biased in adulthood, and gene list of origin indicated the direction of sex-biasing in development.

## Data Availability

All data used for this present study are from publicly available sources. Code for all analyses and results are available at: https://github.com/gabidugan00/Sex_autism_gene_expression.git

https://datadryad.org/dataset/doi:10.5061/dryad.4b8gthtkr#methods

https://www.science.org/doi/10.1126/science.adg5136#supplementary-materials

https://www.science.org/doi/10.1126/science.aea9063#supplementary-materials

## Acknowledgments

We would like to thank the first authors of the papers and respective donors and datasets that these projects rely on, Alex DeCasien, Brie Wamsley, and W. Brad Ruzicka. This research was supported by the Intramural Research Program of the NIMH (ZIAMH002949-08). The Oxford Centre for Integrative Neuroimaging is supported by core funding from the Wellcome Trust (203139//16/Z and 203139/A/16/Z). Computation support for the project was provided by the NIH HPC Biowulf cluster (https://hpc.nih.gov). GD is a DPhil candidate supported by the NIH-Oxford Cambridge Scholars Program. The contributions of the NIH authors are considered Works of the United States Government. The findings and conclusions presented in this paper are those of the authors and do not necessarily reflect the views of the NIH or the U.S. Department of Health and Human Services.

