## Supplementary Materials for "Sex and autism share cell type-specific effects on cortical gene expression"

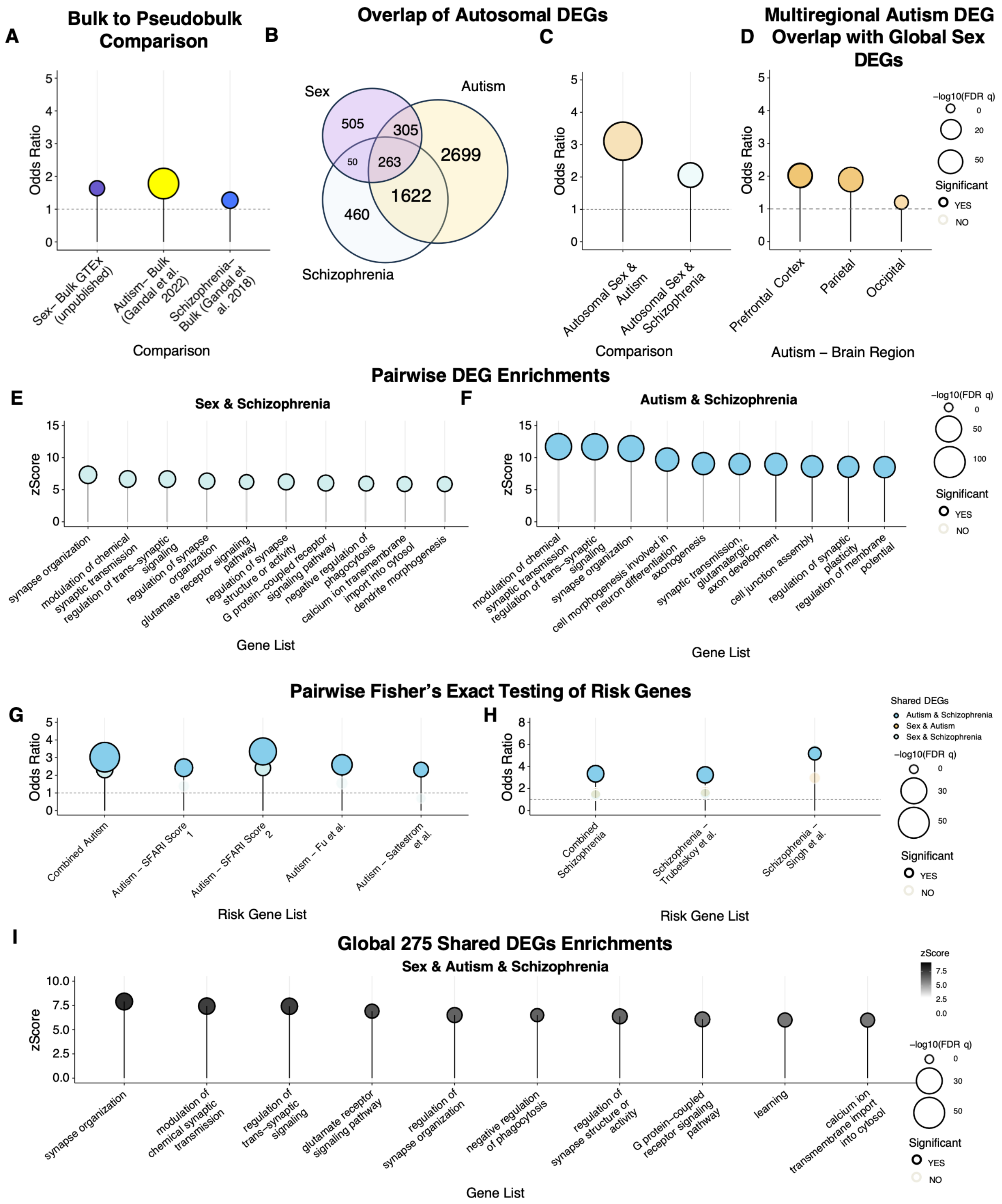


***<u>Figure S1:</u> Supplementary analyses of global DEG overlaps**. (**A**) Odds ratios from Fisher's exact testing to evaluate if the DEGs from bulk-tissue align with the union of DEGs ("global" DEGs) across cell types in snRNA-sequencing. To derive a bulk-multiregional sex DEG list, a joint bulk DEG analysis was performed for sex in GTEX (1) V9 cortical regions (BA9 and BA24) using dream, with individual ID used as a random effect to control for samples originating from the same*

*individual. Prior to dream, outlier samples were removed using PCA and connectivity z-scores, and data was corrected for technical batch effects using RUVr. The bulk autism DEG list was taken from Gandal et al. 2022. (2) and the bulk schizophrenia DEG list was taken from Gandal et al. 2018 (3).* ***(B)*** *Venn diagram summarizing overlap of autosomal shared DEGs identified for sex (4) (purple), autism (5) (yellow), and schizophrenia (6) (blue), with counts indicating shared and unique gene sets.* ***(C)*** *Odds ratios from Fisher's exact testing to evaluate if there is significant overlap of DEGs between sex and autism and sex and schizophrenia, after exclusion of X- and Y-chromosome genes (autosomal).* ***(D)*** *Analysis of overlapping DEGs between global sex-DEG list and autism single-cell dataset across multiple brain regions that were sequenced for smaller sample size (total sample N =12) single-cell RNA-sequencing dataset (2). Odds ratios from Fisher's exact testing of DEGs and background (genes from union of global DEGs in sex across six cortical brain regions and of region-specific DEGs in autism) from single-cell RNA-sequencing across cortical regions. Dot size represents OR; color denotes enrichment significance (FDR q <0.05). We found that the statistically significant DEG overlap was seen for autism DEGs from all three cortical regions. These results show that the transcriptomic overlap observed between autism and sex in our primary analysis is not dependent on sampling of autism DEGs from the frontal cortex - consistent with bulk RNAseq studies suggest that the transcriptional correlates of autism are highly concordant across multiple cortical regions (48).* ***(E–F)*** *Lollipop plots for Gene Ontology (GO) (4) enrichment of shared DEGs from the biological process (BP) ontology, generated using clusterProfiler (5), for* ***(E)*** *sex & schizophrenia shared DEGs and* ***(F)*** *autism & schizophrenia shared DEGs. Enrichment is plotted by zScore (y-axis) and significance (-log10(FDR q), color). Circle size denotes -log10(FDR q) value and color denotes enrichment significance (FDR q <0.05).* ***(G-H)*** *Odds ratios from Fisher's Exact testing of pairwise shared DEGs overrepresentation for independently defined and collated risk gene lists compared to each respective background for* ***(G)*** *autism and* ***(H)*** *schizophrenia risk genes. Dots are colored by shared DEG comparison with light blue (sex and schizophrenia), dark blue (autism and schizophrenia) and golden (sex and autism).* ***(I)*** *GO (7) enrichment of 275 shared DEGs between sex, autism, and schizophrenia global DEG sets shown in Fig. 1A . Enrichment was tested on biological process (BP) ontology gene lists using clusterProfiler (8). Enrichment is plotted by zScore (y-axis) and significance (-log10(FDR q), color). Circle size denotes -log10(FDR q) value and color denotes enrichment significance (FDR q <0.05).*

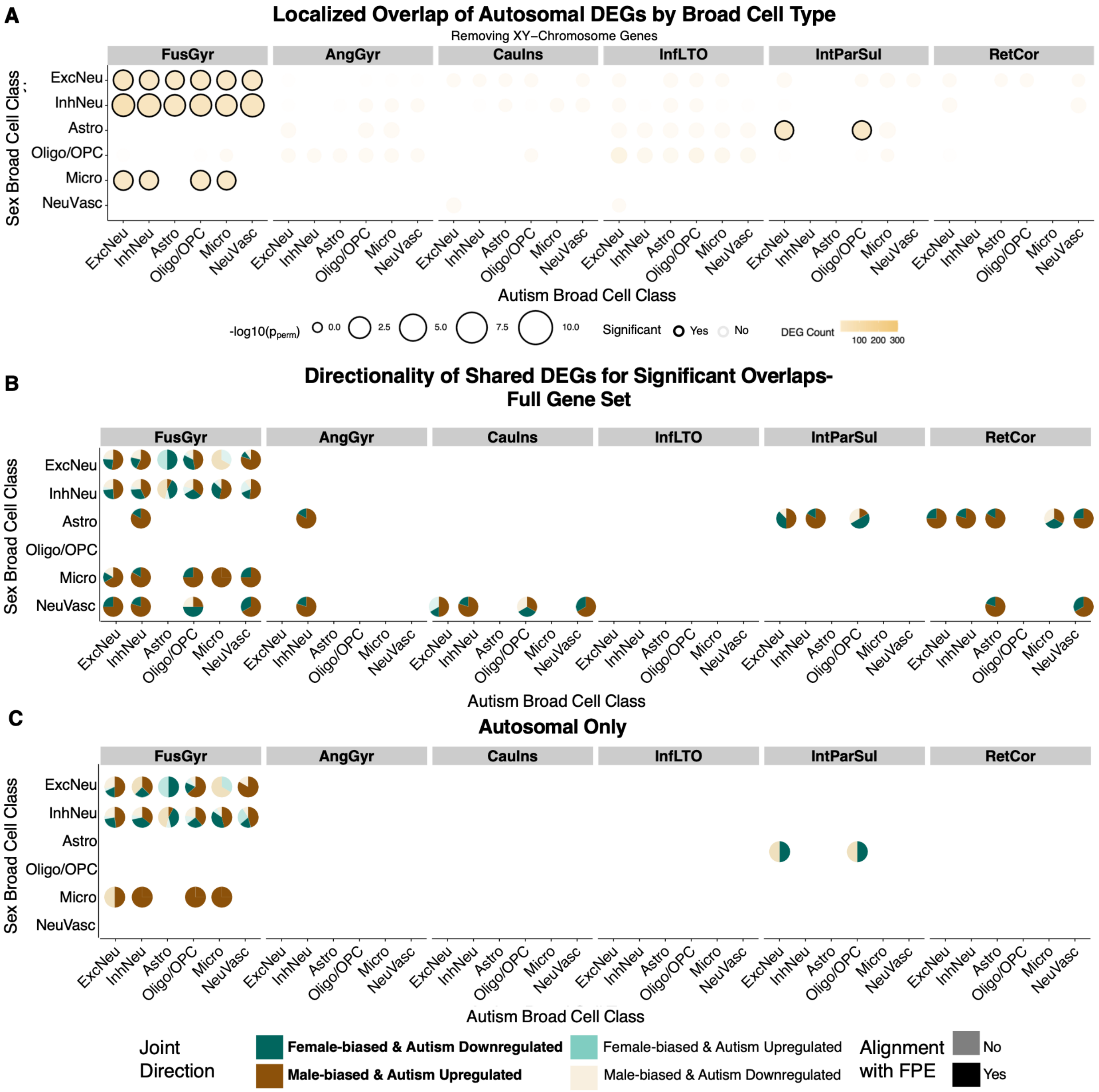


***<u>Figure S2:</u>*** **Supplementary analyses for localized DEG overlaps.** ***(A)*** *Localized overlap testing of autosomal DEGs between autism and sex across brain regions (columns) and broad cell types (rows). Dot size represents overlap size; color denotes enrichment significance (–log10 $p_{perm}$), with outlined points indicating significant overlaps. Significance overlaps were selected and defined by empirical p-value < 0.05 calculated by comparing the number of overlapping DEGs in the original pairwise test to 1000 permutations of sex and recalculated the number of overlapping DEGs for the pairwise comparison.* ***(B)*** *Directionality testing exclusively for significant points of overlap based on permutation testing of sex and comparison to null distribution of overlapping DEGs. Dotplots showing directionality proportion for the shared DEG count within each autism broad cell class (x-axis) and sex brain region (column facet) and cell class (y-axis) pair by direction. Directionality was characterized by mean logFC of shared DEGs by region and broad cell class. Pie charts show coloring considering the direction of gene*

*expression change for both sex and autism: female-biased and autism downregulated (dark teal), male-biased and autism upregulated (dark tan), female-biased and autism upregulated (light teal), and male-biased and autism downregulated (light tan). Male-biased & autism upregulated as well as female-biased and autism downregulated are consistent with a positive correlation (alpha =1) between sex and autism as male >female and autism > control and therefore with theorized female-protective effects. Conversely, the other two direction options are consistent with negative correlation (alpha = 0.5). FusGyr InhNeu (sex) - ExcNeu (autism) overlaps (49% of shared DEGs male-biased and upregulated in autism; FDR q = 0.003).* ***(C)*** *Same as B, with exclusion of non-significant overlaps for directionality tests, but from autosomal shared DEG comparison*

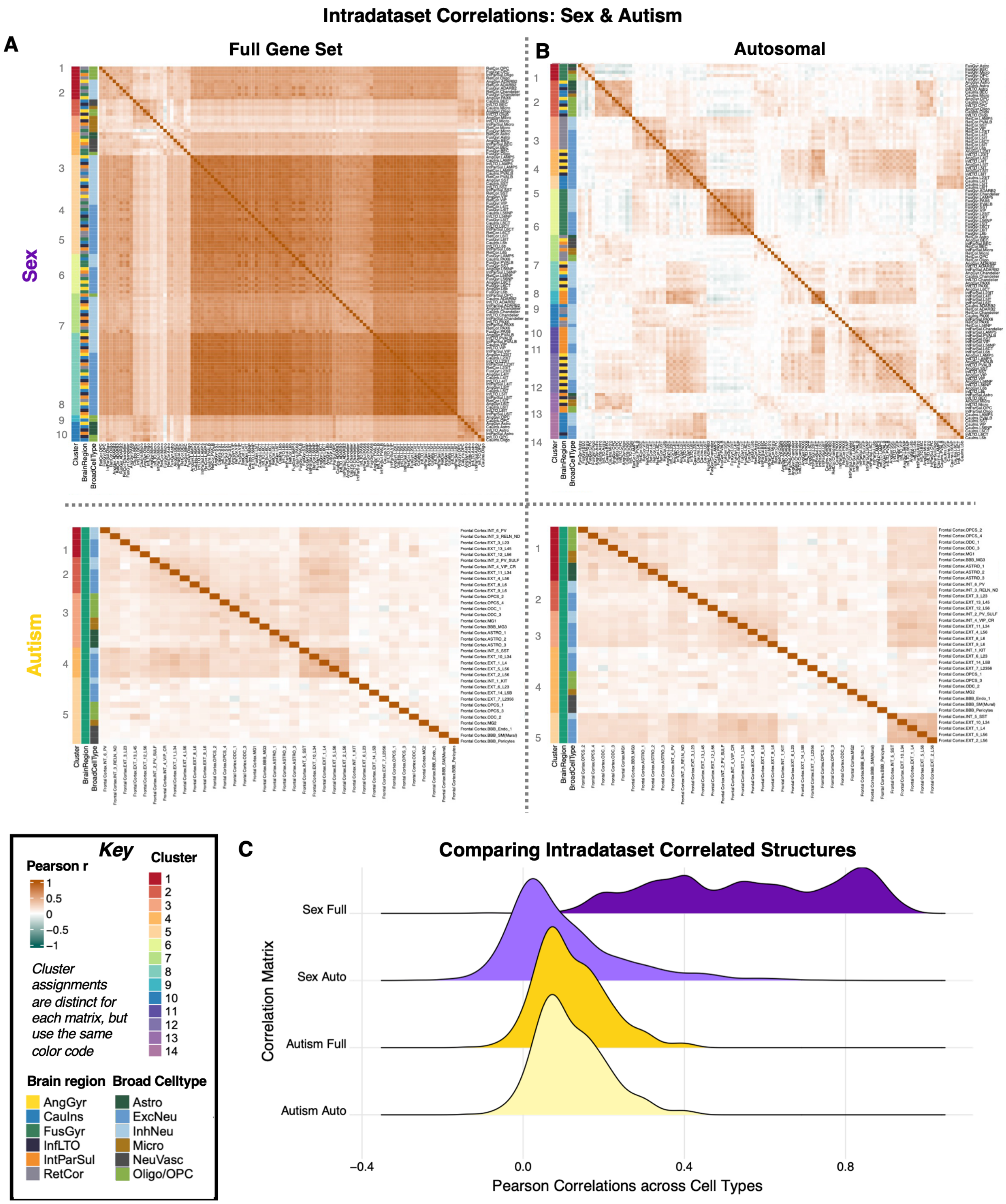


***<u>Figure S3</u>*: Intradataset Pearson correlations in logFC across cell types for sex (4) and autism effects (5)**. *To assess the correlated nature across cell types within the sex and autism datasets, we constructed a brain region - cell type (column) x gene (row) logFC-values matrices for both datasets. For each pairwise combination of cell types, we ran Pearson correlations using the cor function on "pairwise.complete.obs" ensuring correlations were computed only on genes with available logFCs in both cell types. Correlations were run on the full gene set as well*

*as after removal of X- and Y-chromosome genes (autosomal). To test the internal structure across these correlations for sex and autism, we used Weighted Stochastic Block Modeling (WSBM) (31) via the BM_gaussian function and used default parameters to select the optimal fit of block numbers, which maximizes within block similarities. WSBM uses a generative modeling algorithm adaptive for complex datasets to detect blocks as "nodes" based on their correlation relative to the rest of the data.* ***(A)*** *Quadrant of Pearson correlation matrices based on inclusion of sex-chromosome genes and dataset tested. Rows and columns of the matrices are organized by clusters determined by WSBM (9). The number and composition of the blocks changed based on the input matrices, but the color scale was constant.* ***(B)*** *Same as A but from autosomal shared DEG comparison. We observed significantly stronger intradataset correlations within the sex-biased dataset (mean r = 0.56) compared to the autism-associated dataset (mean r = 0.14). The disparity in intradataset correlations was largely driven by sex-chromosome dosage: as expected, excluding sex-chromosome genes had a minimal impact on autism correlations but a substantial effect on the sex correlations, where the mean correlation coefficient dropped from 0.56 to 0.10. Upon removing sex-chromosome genes, the blocking in sex was driven more by brain region than cell type, indicative of the autosomal effects of sex being organized more by neuroanatomy.* ***(C)*** *Ridge plot of the distribution of Pearson correlation values across celltypes for each correlated matrix: sex full, sex, auto, autism full, autism auto.*

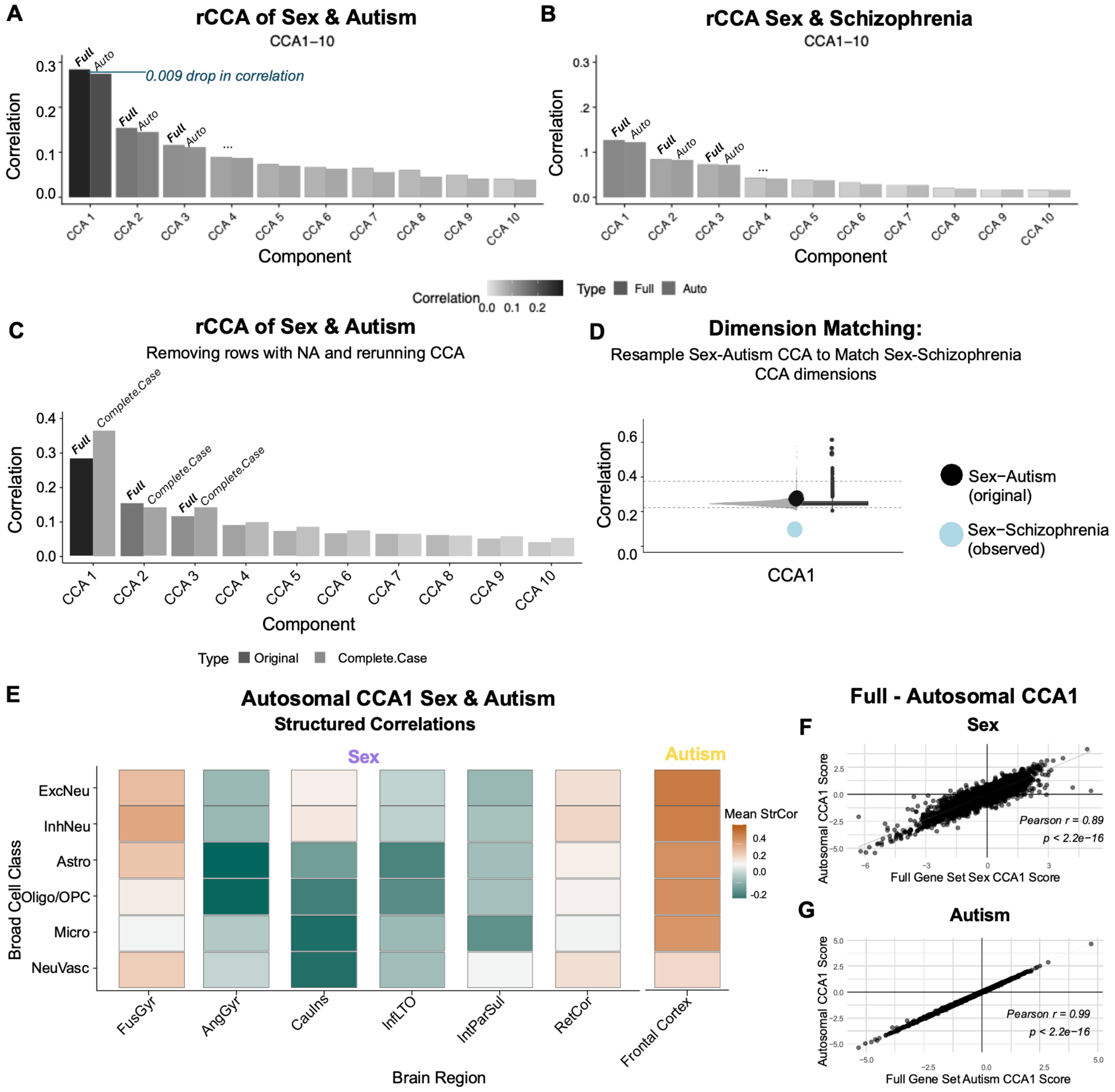


***<u>Figure S4:</u> Sensitivity tests for regularized CCA (rCCA) for Sex & Autism and Sex & Schizophrenia. (A-B)*** *Barplot showing rCCA results comparing full (black) to autosomal (grey) CCA results for Sex & Autism **(A)** and Sex & Schizophrenia **(B)**. Sex-chromosome genes were removed from the input gene expression matrices and rCCA was rerun to derive the autosomal CCAs. **(C)** Barplot showing Sex-Autism CCA results comparing full (black) to complete case (grey) CCA results. Complete case rCCA describes rCCA run on the matrices after removing any genes that had NA values in either matrix (n=3749 genes). **(D)** Violin plot showing distribution of CCA1 correlation magnitudes from 1000 resamplings of autism matrix to align the dimensions of the Sex-Autism CCA and Sex-Schizophrenia CCA. The black dot indicates the CCA1 magnitude of the original Sex-Autism CCA1, and the blue dot indicates the CCA1 magnitude of the observed Sex-Schizophrenia CCA1. The dashed grey lines show the*

*2.5th and 97.5th percentiles of the 1,000 resampled Sex-Autism CCA1 magnitudes (the 95% percentile interval). Importantly, the observed Sex-Schizophrenia CCA1 correlation magnitude is completely outside of the distribution of the resampled Sex-Autism tests, indicating that the lower value is not a result of different dimensions.* ***(E)*** *Average Pearson correlation between logFC and the first canonical component (Autosomal CCA1) of sex and autism after excluding X- and Y-chromosome genes per broad cell type, calculated across all genes (i.e. rCCA Structure Correlations).* ***(F-G)*** *Correlation plots of CCA1 weights for* ***(F)*** *sex and* ***(G)*** *autism between full and autosomal CCA1 scores.*

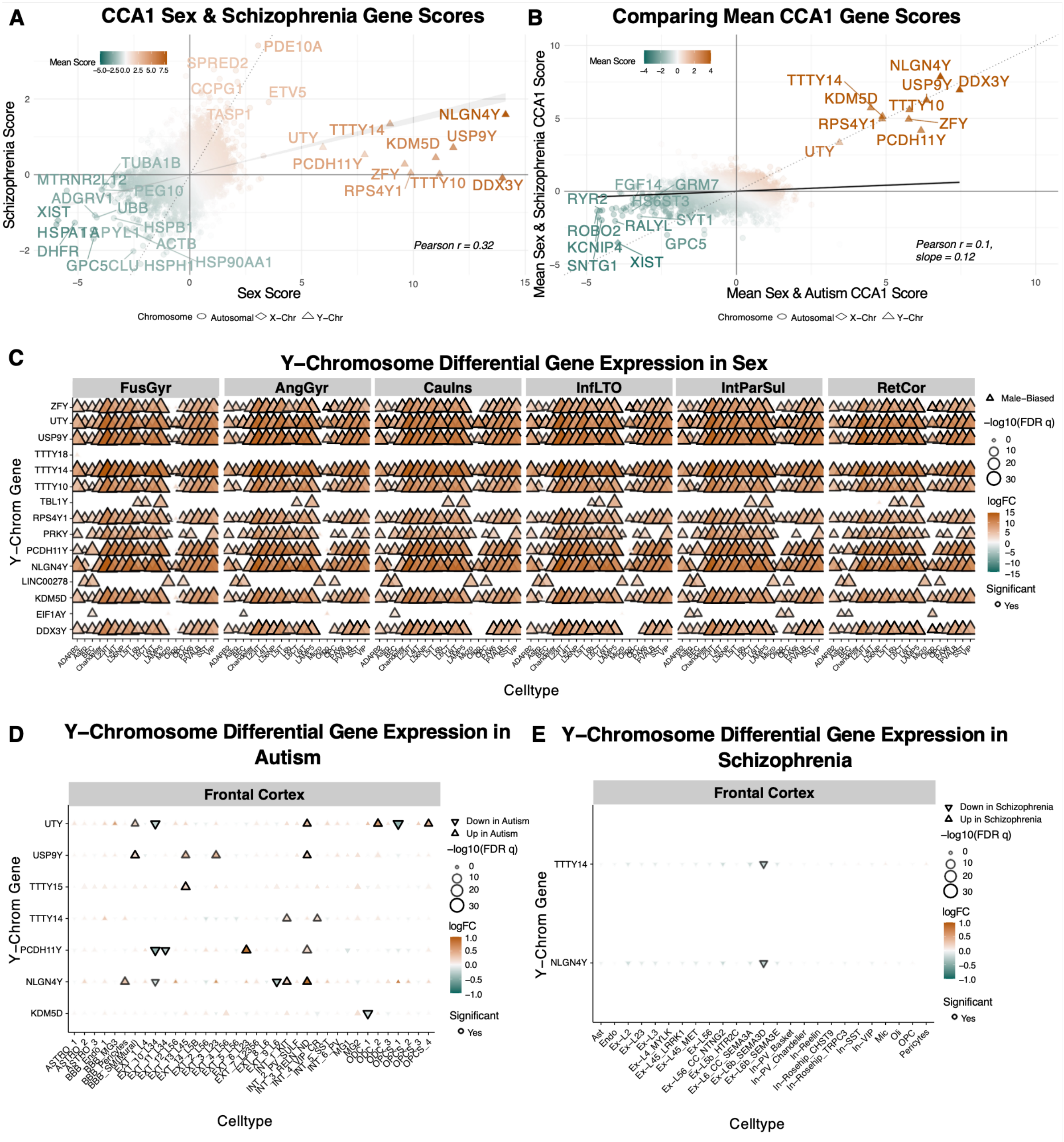


*Inclusion criteria for sex-chromosome genes was that they must meet differential expression thresholds in at least one cell type*

***Figure S5:*** ***Comparing CCA1 for Sex & Autism and Sex & Schizophrenia: (A)*** *Scatter plot of CCA1 gene scores for sex (x-axis) and schizophrenia (y-axis). Each point represents a single gene and the top 30 genes by |mean score| of sex and schizophrenia are labeled with text.* ***(B)*** *Scatter plot of mean gene scores for CCA1 comparing Sex & Autism (x-axis) and Sex & Schizophrenia (y-axis). Each point is a gene; color denotes mean score. Pearson correlation r values are labeled, as well as the slope of y~x. Most sex-chromosome genes sit along or close to a slope of 1, indicating comparable scoring of these genes in both CCA1s. (****C,D,E****) Dot plots showing differential gene expression (DGE) for Y-chromosome genes in* ***(C)*** *sex (4)* ***(D)*** *autism (5)* ***(E)*** *schizophrenia (6). Each dot represents a gene–cell type pair; color denotes logFC, and* *dot size reflects –log10(FDR q). Black outlines denote statistically significant effects. Genes were included if they were differentially expressed in at least one cell type*

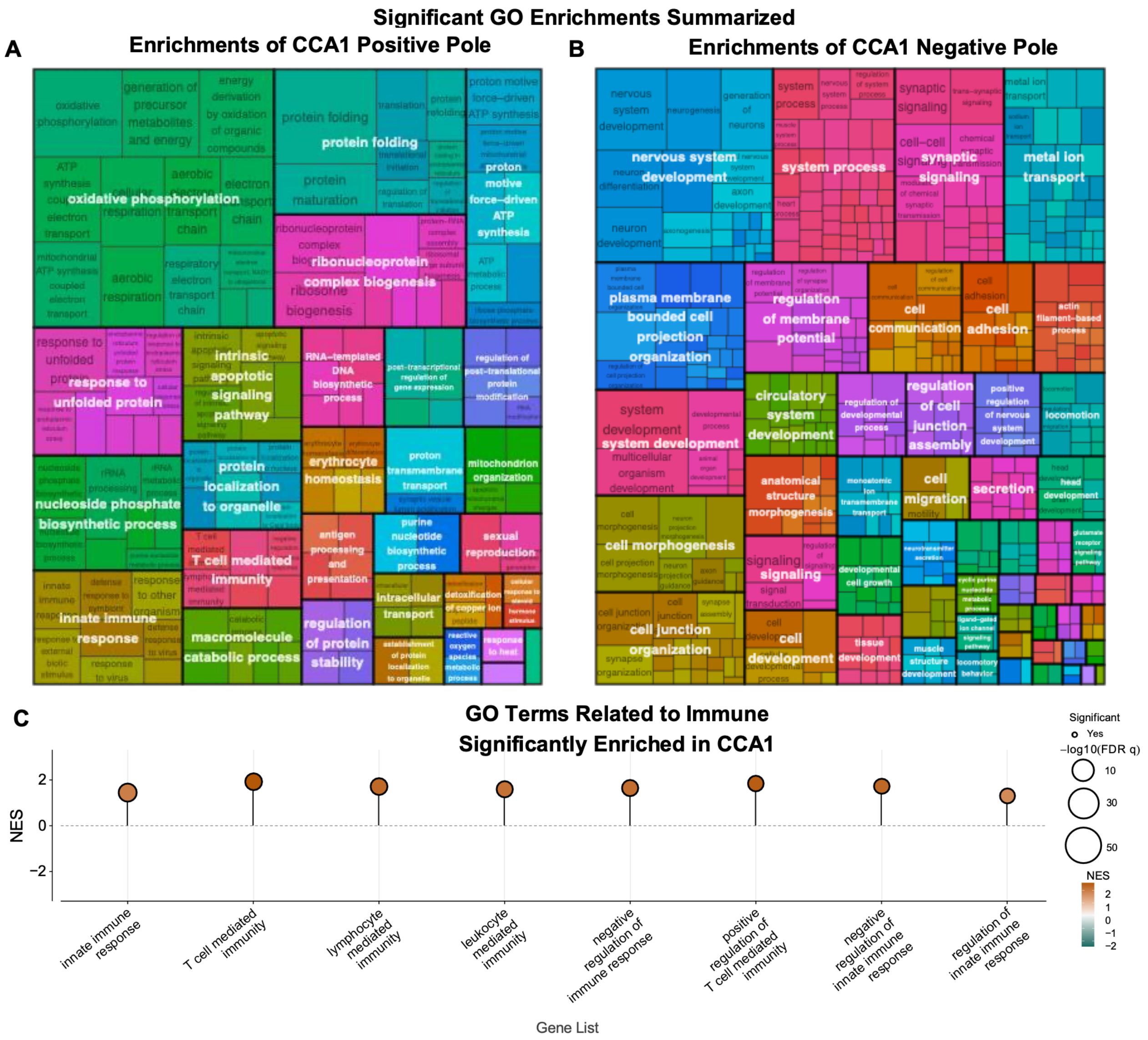


***<u>Figure S6:</u> GO enrichments for the poles of CCA1:*** *rrvgo (10) treemap plots of significant (FDR q <0.05) GO BP terms for the positive* ***(A)*** *negative* ***(B)*** *poles of CCA1.* ***(C)*** *Lollipop plot of selected immune gene list enrichment results from fGSEA of full CCA1 gene scores. Terms were selected if they included "immun" in the GO pathway name and were significant (FDR q <0.05).*

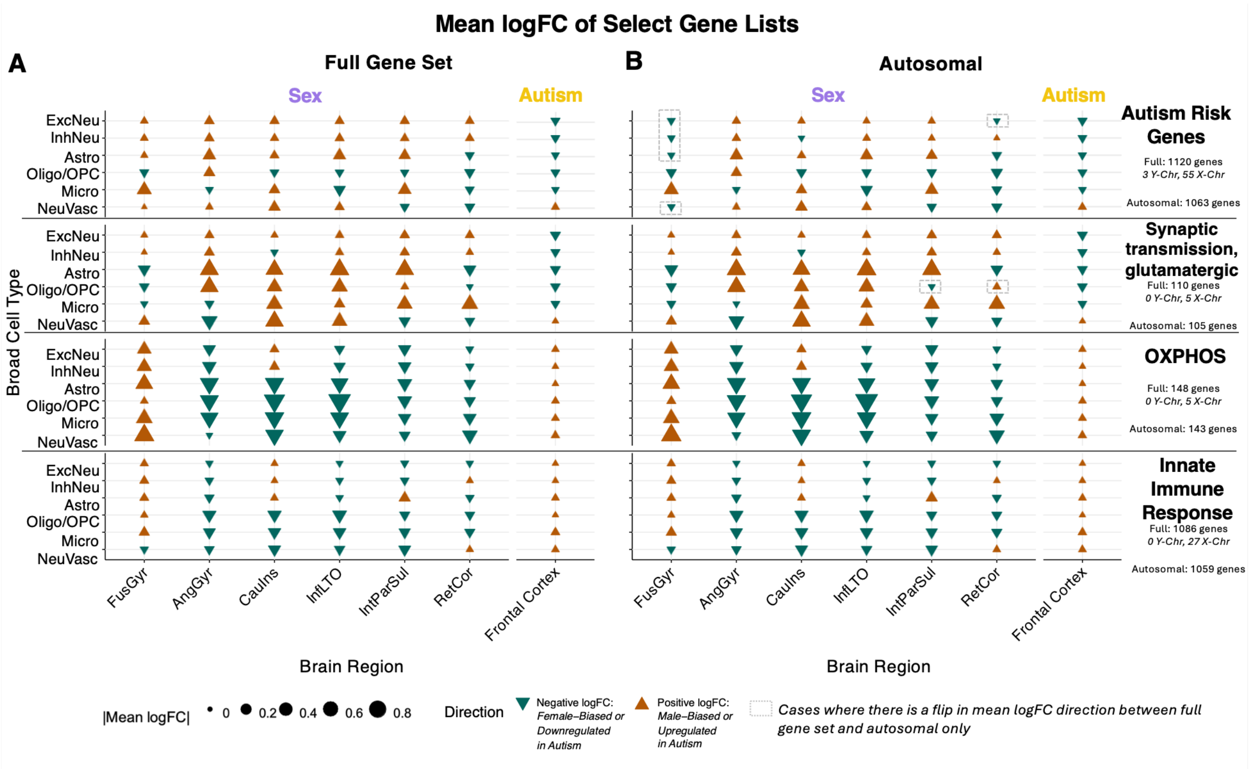


***<u>Figure S7:</u> Relating enriched annotations from the sex & autism CCA1 to observed cell class gene expression changes for sex and autism.*** ***(A)*** *Dotplots of the mean logFC of select gene lists to project the annotation enrichments for CCA1 back to the gene expression changes observed for sex and autism. Gene sets were chosen to represent: the combined autism risk gene set (n = 1,120 genes); the most negatively enriched GO term in CCA1 — synaptic transmission, glutamatergic (n = 110 genes); the most positively enriched GO term in CCA1 — OXPHOS (n = 148 genes); and the most positively enriched immune-related GO term — innate immune response (n = 1,086 genes). Color and shape denote the direction of the mean logFC (across genes and fine cell types within the broad categorization) showing positive logFC - male-biased or upregulated in autism (upward facing triangle; dark tan) and negative logFC - female-biased or downregulated in autism (downward facing triangle; dark teal).* ***(B)*** *As in A, but after removal of X- and Y-chromosome genes (autosomal).*

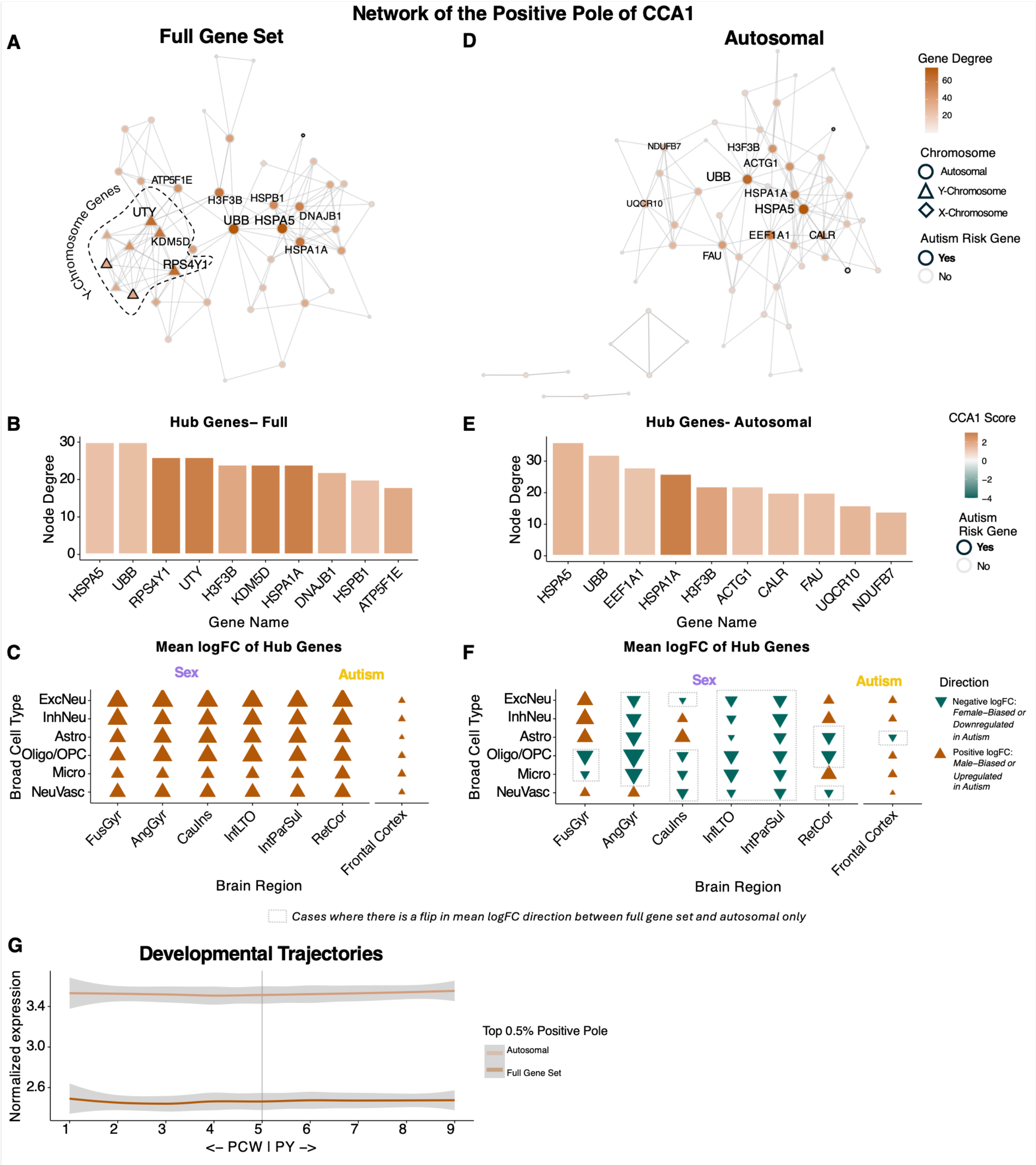


***<u>Figure S8:</u> Biological characterization of the positive pole of the sex-autism CCA1 axis.*** ***(A)*** *Protein-protein interaction (PPI) networks from STRINGdb (11) for the top 0.5% of the positive pole.* ***(B)*** *Barplots of the top 10 most connected genes (x-axis) based on number of connections, "Node Degree" (y-axis). Bars are outlined if the gene is in the collated autism risk gene list for the* ***(C)*** *Dotplots of the mean logFC of hub genes to project the CCA1 directions back to the gene expression changes observed for sex and autism. Color and shape denote the direction of the mean logFC (across hub genes and fine cell types within the broad categorization) showing positive (upward facing triangle; dark tan) and negative (downward facing triangle; dark teal). The presence of Y-chromosome genes (KDM5D, UTY, and RPS4Y1)*

*led to a ubiquitous positivity of the mean logFCs. This motivated an analysis after removing X- and Y-chromosome genes.* ***(D-F)*** *As in A-C, but after removal of X- and Y-chromosome genes (autosomal). Now, we observe male-biasing of positive network hub genes for FusGyr neurons and female-biasing for AngGyr glial cells, consistent with structured correlations* ***(F). (G)*** *Developmental trajectory curves from PsychENCODE (12) of normalized gene expression for full gene set and autosomal positive pole of CCA1 across 10 developmental period: period 1: post-conceptional week (PCW) 5-9, period 2: PCW 12-13, period 3: PCW 16-18, period 4: PCW 19-22, period 5: PCW 35- PY 0.3, period 6: post-natal year (PY) 0.5-2.6, period 7: PY 2.8-10.7, period 8: PY 13-19, period 9: PY 21-64. The shaded region indicates 95% confidence intervals.*

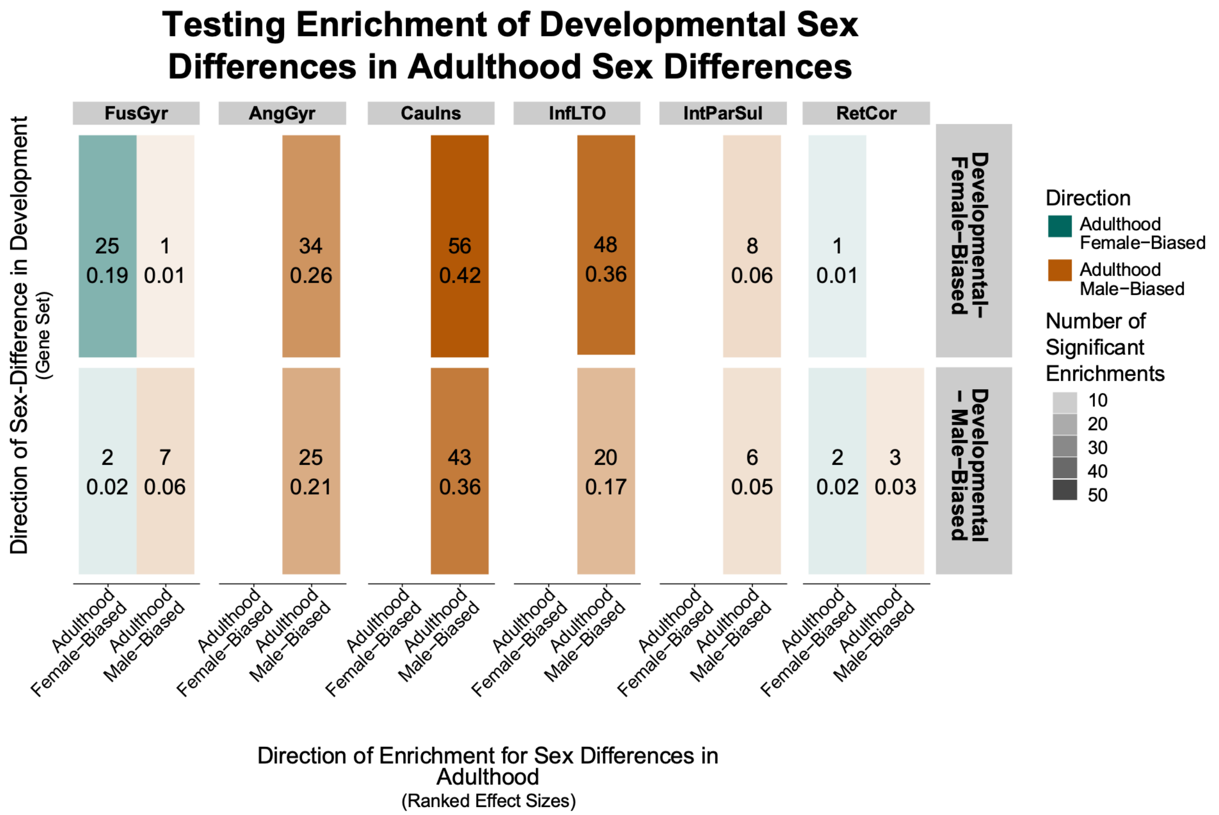


***<u>Figure S9:</u> Testing continuity of transcriptomic sex-differences across development to adulthood.*** *We used rank-based analyses to test if sets of significantly sex-biased genes in a developmental cortical dataset (13) show enrichments in the ranking of genes by sex logFC in our adult cortical dataset (4). There are 120 developmental-male-biased gene sets and 132 developmental female-biased gene sets across cell types. We tested for enrichment across these gene sets within the ranked mean logFCs for each of six broad cell class in each of the six brain regions. The first number in each square shows the counts, which represents the number of DEG lists within each class that show a significant rank-based enrichment, the second number shows the proportion, which represents the ratio of significant enrichments compared to the number of gene lists tested for that category, e.g. 25 significantly downregulated (female-biased in adulthood) enrichments for the 132 developmental female-biased gene lists: 25/132 = 0.19, and color denotes the direction of that enrichment [negative NES indicating female-biased in adulthood (teal) and positive NES indicating male-biased in adulthood (tan)]. The upper left and bottom right quadrant for each brain region show sex-differences that are consistent between development and adulthood, e.g. female-biased DEGs in development enriched for female-biased genes in adulthood (top quadrant).*

### Supplementary Table Legends:

**<u>Table S1:</u> Relevant information for overview of sex, autism, and schizophrenia datasets,** including reporting number of individuals, number of males and females, quality control metrics, and differential gene expression model derived from original analyses.

**<u>Table S2:</u> Differential genes expression (A)** Gene-level characterization of differential expression from sex (4), autism (5), and schizophrenia (6) datasets, noting whether each gene was tested in the dataset and whether it met the significance threshold for classification as a DEG. **(B)** Fisher's exact testing output for bulk to pseudobulk comparisons. For autism, bulk DEG lists were taken from Gandal et al. 2022 (2), which was a multiple cortical region bulkRNAseq analyses. For schizophrenia, bulk DEG lists were taken from Gandal et al. 2018 (3) a bulkRNAseq analysis using PsychENCODE (14) RNAseq. **(C)** Fisher's exact test inputs and outputs for localized shared DEG test by autism cell class and sex region-by-cell class**. (D)** Fisher's exact test input and outputs for testing direction of localized DEGs in autism and sex for each significant (pemp <0.05) cell class pair.

**<u>Table S3</u>: Gene score outputs from CCA1** for **(A)** sex and autism **(B)** sex and schizophrenia, with sex-chromosome genes and excluding sex-chromosome genes (autosomal).

**<u>Table S4</u>: Enrichment testing of CCA1 results** across Gene Ontology Biological Process (7) and risk gene set terms. **(A)** Mean Sex-Autism CCA1 scores **(B)** Sex scores from Sex-Autism CCA1 **(C)** Autism scores from Sex-Autism CCA1 **(D)** Sex scores from Sex-Schizophrenia CCA1 **(E)** Schizophrenia scores from Sex-Schizophrenia CCA1.
